# Urbanization exacerbates age-associated declines in cardiometabolic health in Turkana and Orang Asli

**DOI:** 10.1101/2025.06.06.25329160

**Authors:** Marina M. Watowich, Layla Brassington, Amy Longtin, Selina Wang, Ryan Rossow, Kathleen D. Reinhardt, Echwa John, John C. Kahumbu, Patricia Kinyua, Anjelina Lopurudoi, Francis Lotukoi, Charles Miano, Benjamin Muhoya, Boniface Mukoma, Tan Bee Ting A/P Tan Boon Huat, Kar Lye Tam, Yvonne A L Lim, Dino Martins, Sospeter Ngoci Njeru, Kee Seong Ng, Vivek V. Venkataraman, Ian J. Wallace, Julien F. Ayroles, Michael Gurven, Thomas S. Kraft, Amanda J. Lea

**Affiliations:** Department of Biological Sciences, Vanderbilt University, Nashville, TN, USA, 37232; Department of Anthropology and Archaeology, University of Calgary, Calgary, Alberta, Canada T3B 5C1; Turkana Health and Genomics Project, Kenya Medical Research Institute, Nairobi, Kenya 54840-00200; Center for Community Driven Research, Kenya Medical Research Institute, Nairobi, Kenya 54840-00200; Lewis-Sigler Institute for Integrative Genomics, Princeton University, Princeton, New Jersey, USA 08540; Department of Ecology and Evolutionary Biology, Princeton University, Princeton, New Jersey, USA 08540; Department of Parasitology, Universiti Malaya, Kuala Lumpur, Malaysia 50603; Turkana Basin Institute, Stony Brook University, Stony Brook, NY 11794; Department of Medicine, Universiti Malaya, Kuala Lumpur, Malaysia 50603; Department of Anthropology, University of New Mexico, Albuquerque, New Mexico, USA 87131; Department of Anthropology, University of California Santa Barbara, Santa Barbara, USA 93106; Department of Anthropology, University of Utah, Salt Lake City, Utah, USA 84112; Evolutionary Studies Initiative, Vanderbilt University, Nashville, TN, USA 37232; Vanderbilt Genetics Institute, Vanderbilt University, Nashville, TN, USA 37232

## Abstract

Declines in cardiometabolic health among older individuals are so ubiquitous in Western, high-income countries that non-communicable diseases (NCDs) like type 2 diabetes, hypertension, and cardiovascular disease have been termed “diseases of aging”. In contrast, research from non-industrial contexts has found low rates of cardiometabolic NCDs in old age, suggesting protective effects of lifestyle. To test if industrialization and urbanization generates or magnifies age-associated cardiometabolic health patterns, within-population analyses are needed. We worked with Turkana pastoralists of Kenya and Orang Asli mixed subsistence groups of Peninsular Malaysia—two groups that are transitioning from non-industrial to urban, market-integrated lifestyles. We find that rural, non-industrial environments produce minimal to modest age-dependent increases in body size, lipid, and blood pressure traits, and that urban environments significantly amplify age effects in repeatable ways across two distinct populations. However, we did not find that urban environments consistently accelerate biomarkers of more generalized functional capacity and biological aging, namely grip strength, walking speed, and epigenetic age. Together, these findings challenge the view that cardiometabolic “diseases of aging” are an intrinsic feature of aging, instead implicating urban lifestyle features as drivers of age-associated variation; however, these same lifestyle exposures may have heterogeneous effects on biological aging. These results underscore the urgency of understanding how rapid lifestyle changes shape aging trajectories, especially in populations undergoing industrial transitions.

## Introduction

Increased chronological age is the single greatest risk factor for most of the non-communicable diseases (NCDs) that burden societies globally, including ischemic heart disease, stroke, diabetes, neurodegenerative disorders, kidney and liver disease, osteoarthritis, and many cancers (1–4). These conditions are commonly classified as “diseases of aging” (5, 6) and are a well-established feature of high-income, industrialized human societies; this conclusion is supported by both longitudinal and cross-sectional studies, leading to the suggestion that age-associated declines are driven by the normative human aging process and are near-universal byproduct of getting older (7–12). At the biological level, age-related increases in morbidity and mortality are expected to be driven by reduced somatic maintenance and repair capacity, which is thought to be an unavoidable evolutionary byproduct of relaxed selection following first reproduction (5, 13–21).

Relative to most of human evolutionary history and to modern day non-industrial populations (e.g., hunter-gatherers, horticulturalists, pastoralists), industrialized societies exhibit low levels of physical activity, diets high in processed foods, more extreme social inequality, disturbed sleep, and high exposure to environmental toxins—all known risk factors for cardiometabolic disease—making it challenging to disentangle the effects of normative human aging from cumulative urban lifestyle exposure (22–24). Mounting evidence from non-industrial groups suggests these populations exhibit minimal age-related differences in cardiometabolic biomarkers—including coronary artery calcification, blood pressure, serum cholesterol, and body fat—and exhibit low overall prevalence of cardiometabolic NCDs, despite vast diversity in diets and subsistence strategies (25–35). However, most previous studies that have investigated how lifestyle modifies cardiometabolic age effects have focused on cross population comparisons (e.g., non-industrial groups versus the US) which unavoidably confound genetic background with lifestyle. A more powerful approach is to study the effects of lifestyle variation within populations. Yet, previous within-population studies have been very small, population-specific, or focused on one or a few outcome measures. Additionally, very little work has investigated molecular or physiological biomarkers of the aging process itself—such as epigenetic dysregulation, physical function, or frailty—in non-industrial groups (36). Thus, it remains uncertain whether urban, industrialized lifestyle exposures impact fundamental biological aging processes throughout the body, or instead, if their effects are limited to specific physiological systems and outcomes (e.g., biomarkers of cardiometabolic health) (37–39).

Here we 1) test the hypothesis that more industrialized, urban, and market-integrated lifestyles exacerbate age effects on cardiometabolic health and 2) investigate the extent to which these effects are recapitulated in general biological markers of physiological decline. To do so, we worked with Turkana pastoralists of Northwest Kenya and the Orang Asli of Peninsular Malaysia, Indigenous groups that have been experiencing rapid and intense lifestyle change over the last few generations. These groups thus provide a rare opportunity to test how urbanized, market-integrated lifestyles accelerate age-related declines in the absence of major confounds between genetics and environment (34, 40–43). Using cross-sectional data from 3734 Turkana and 956 Orang Asli, we tested the extent to which 16 biomarkers of cardiometabolic health differed throughout the lifespan between Turkana and Orang Asli exposed to non-industrialized, subsistence-based versus market-integrated, urban lifestyles (44, 45) (Figures 1A-B, S1). We investigated the degree to which these age by lifestyle patterns were sex-dependent, and compared them to publicly available data from people living in the US (46). We also tested whether urban environments influenced age-related differences in established biomarkers of the generalized aging process: 1) grip strength and walking speed, which reflect physiological and functional decline (47, 48) and 2) epigenetic age acceleration estimated from DNA methylation data (40, 49–51). Our findings support the conclusions that cardiometabolic ‘diseases of aging’ are not an inevitable part of human aging, that age-associations of cardiometabolic outcomes are highly environmentally modifiable, and that these health declines can unfold independently from those of more generalized biological mechanisms of aging.

**Figure 1:**
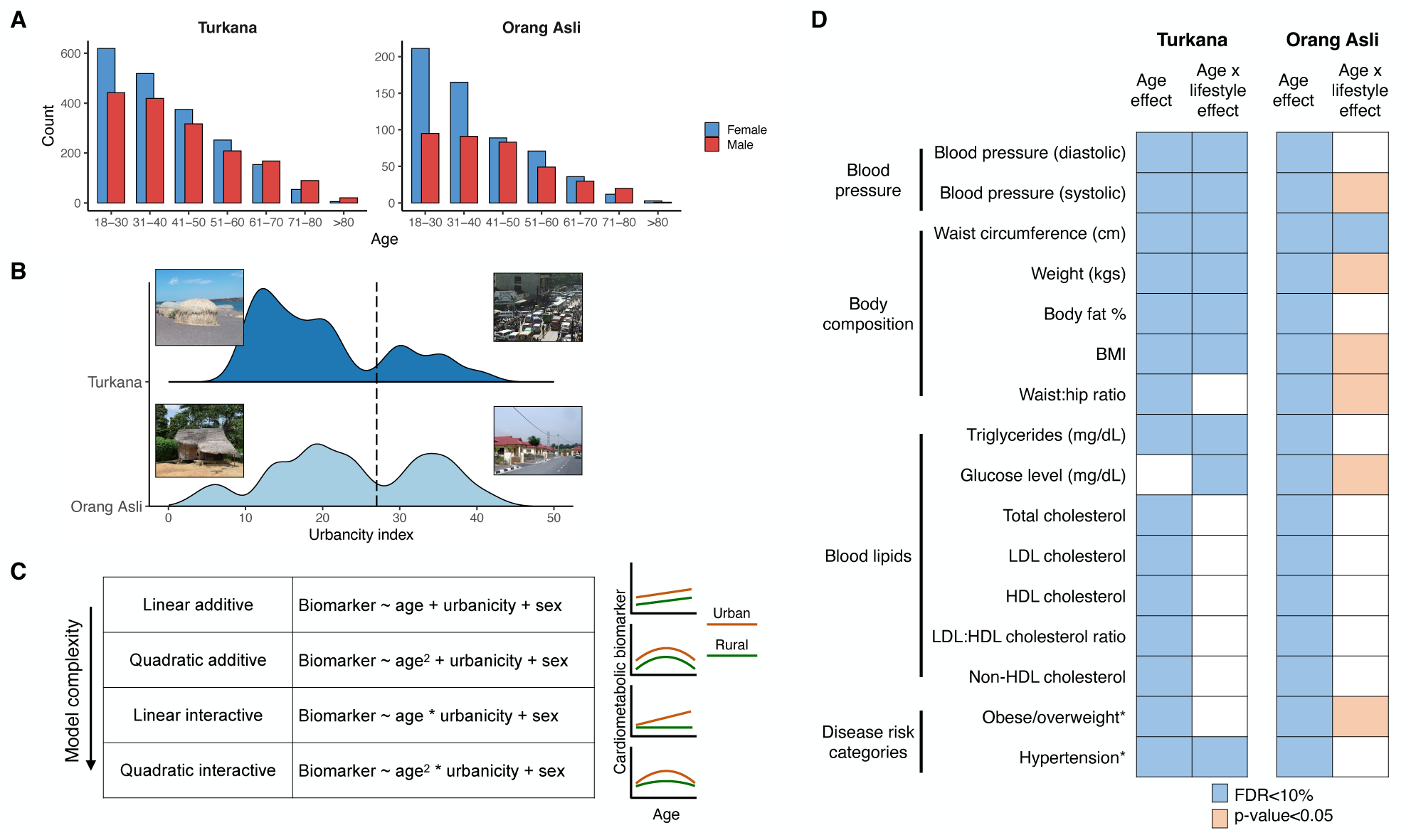
Study overview and summary of main effects. (A) Age and sex distributions of the Turkana and Orang Asli datasets (further detailed in Table S1). (B) Location-based measure of urbanicity for each sampling location in Kenya (Turkana) and Malaysia (Orang Asli). A higher urbanicity score indicates the location has more industrialized infrastructure, market access, acculturation, and higher population density. Photos show representative ‘low’ and ‘high’ urbanicity locations in each country. The black dotted line shows the cut-off used to differentiate between ‘rural’ and ‘urban’ individuals for plotting purposes. The table in (C) shows the models compared for within-Turkana analyses to establish which age term best fit the data for each biomarker. (D) Outcomes of main statistical analyses from the Turkana and Orang Asli dataset, with cardiometabolic biomarkers in columns. Categorical outcomes are denoted by asterisks. Focusing on the best fit models from our model selection approach, we asked which biomarkers had main effects of age and age x lifestyle effects. The significance threshold at which the effect was detected is shown via shading color. FDR refers to the false discovery rate following multiple hypothesis testing correction.

## Results

### Age effects on cardiometabolic health are exacerbated by urban lifestyles

To characterize how lifestyle moderates age effects (i.e., age-related slopes) on cardiometabolic health in a design that minimally confounds genetic background, environment and culture, we drew on data from two anthropological projects with long-standing community relationships: the Turkana Health and Genomics Project (3734 participants spanning 63 locations) and the Orang Asli Health and Lifeways Project (956 participants spanning 29 locations). Both of these projects have collected extensive demographic, interview, and biomarker data using very similar methods, and are designed to work with communities that span a lifestyle gradient from traditional, subsistence-level lifestyles to fully urban and industrialized contexts (42, 43). Lifestyle transitions in Kenya and Malaysia are composed of intertwined processes including market integration (participation in the cash economy), acculturation (assimilation to the dominant culture), industrialization (transition to a manufacturing-based economy), and urbanization (proximity to and interaction with urban infrastructure) (52).

The Turkana people have traditionally practiced nomadic pastoralism; however, in the last few generations, many Turkana have moved to urban centers due to ongoing socioeconomic development and industrialization of Kenya as well as increasingly frequent droughts that threaten livestock (42, 53). ‘Orang Asli’ is a term that broadly refers to the Indigenous peoples of Peninsular Malaysia but encompasses 19 distinct ethnolinguistic groups with varied subsistence histories including hunting, gathering, foraging, and subsistence-level agriculture (43, 54). Orang Asli ethnolinguistic groups are more closely genetically similar to one another than other South East Asian populations. All Orang Asli, to varying degrees, have recently experienced acculturation, market-integration, and urbanization driven largely by deforestation, resource extraction, and government resettlement schemes (54–57). Previous work by our group has 1) validated a continuous index of “urbanicity” that captures exposure to urban infrastructure and can be calculated from comparable data in both populations (Figures 1B, S1) (44, 45); and 2) shown that both Orang Asli and Turkana living in more urban areas have poorer overall cardiometabolic health (42, 45).

Building on these comparable datasets and lifestyle gradients, we first tested the extent to which Turkana and Orang Asli exhibited age, lifestyle (i.e., “urbanicity”), and age x lifestyle effects for 16 cardiometabolic phenotypes (Figure 1C-D; distributions of biomarkers Figure S2-3). All models controlled for sex and compared the fit of both linear and quadratic age terms in each population, following previous work (12, 25, 32). In both groups, we found that essentially all biomarkers exhibited some type of age effect in the overall dataset (the only exception was glucose levels in Turkana; 10% false discovery rate, FDR (Figures 1D, 2A-D, S4; Tables S2-3).

These trends broadly fit expected patterns and we observed that quadratic curves fit age-related patterns better than linear patterns for many biomarkers (13 of 16 in both populations), with many biomarkers peaking among middle-aged individuals and decreasing in older individuals. Of the 16 biomarkers measured in Turkana, we found that 9 biomarkers exhibited an age x lifestyle effect (10% FDR). In Orang Asli, for which we had a smaller sample size, we found that 1 biomarker exhibited an age x lifestyle effect at a stringent threshold (10% FDR) and 6 biomarkers exhibited an age x lifestyle effect at a less stringent threshold (p < 0.05). Consistent with our previous work, we also observed pervasive effects of lifestyle on cardiometabolic biomarkers, with 13 of 16 biomarkers significantly associated with lifestyle in both datasets (10% FDR).

All cardiometabolic health biomarkers with significant age x lifestyle interactions followed patterns we predicted from previous cross-population studies (58, 59): individuals exposed to more urban environments exhibited stronger age effects that were generally indicative of poorer cardiometabolic health (exacerbated blood pressure, LDL and total cholesterol, adiposity), while age had much more subtle impacts on health in rural settings. This was true for all 9 biomarkers with significant age x lifestyle interactions in Turkana (FDR < 10%): waist circumference, body fat percentage, BMI, weight, triglycerides, systolic and diastolic blood pressure, blood glucose, and hypertension (Figures 1D, 2A-C, S4, S5; Table S2). To confirm whether age effects are minimal to nonexistent in fully non-industrial settings, we also tested for age effects in a subset of Turkana individuals subsisting primarily from pastoralism and with minimal evidence of market integration (n = 330); here, we only found evidence that four biomarkers showed significant differences with age: body fat percentage was higher in older individuals, while weight, BMI, and HDL cholesterol were lower in older individuals (FDR < 10%; Figures S4, S6; Table S4).

We observed similar exacerbation of age-associated cardiometabolic disease in the Orang Asli dataset (n = 956; Figures 2D, S4, S7). Specifically, we found that waist circumference exhibited significantly amplified age effects in urban environments (FDR < 10%), and that BMI and obesity/overweight exhibited similar patterns at a more liberal significance threshold (p < 0.05; Figures 2D, S4, S7; Table S3). Importantly, when we compared the effect sizes of the age x lifestyle interaction effects in the Orang Asli to those estimated in the Turkana, we found very strong concordance (Pearson’s r = 0.7, p = 2.72 x 10^−3^; Figure 2E). This degree of generalizability is remarkable given that these groups live on separate continents, in very distinct ecologies, and the lifestyle transition is occurring for different reasons and in different ways in both populations.

**Figure 2:**
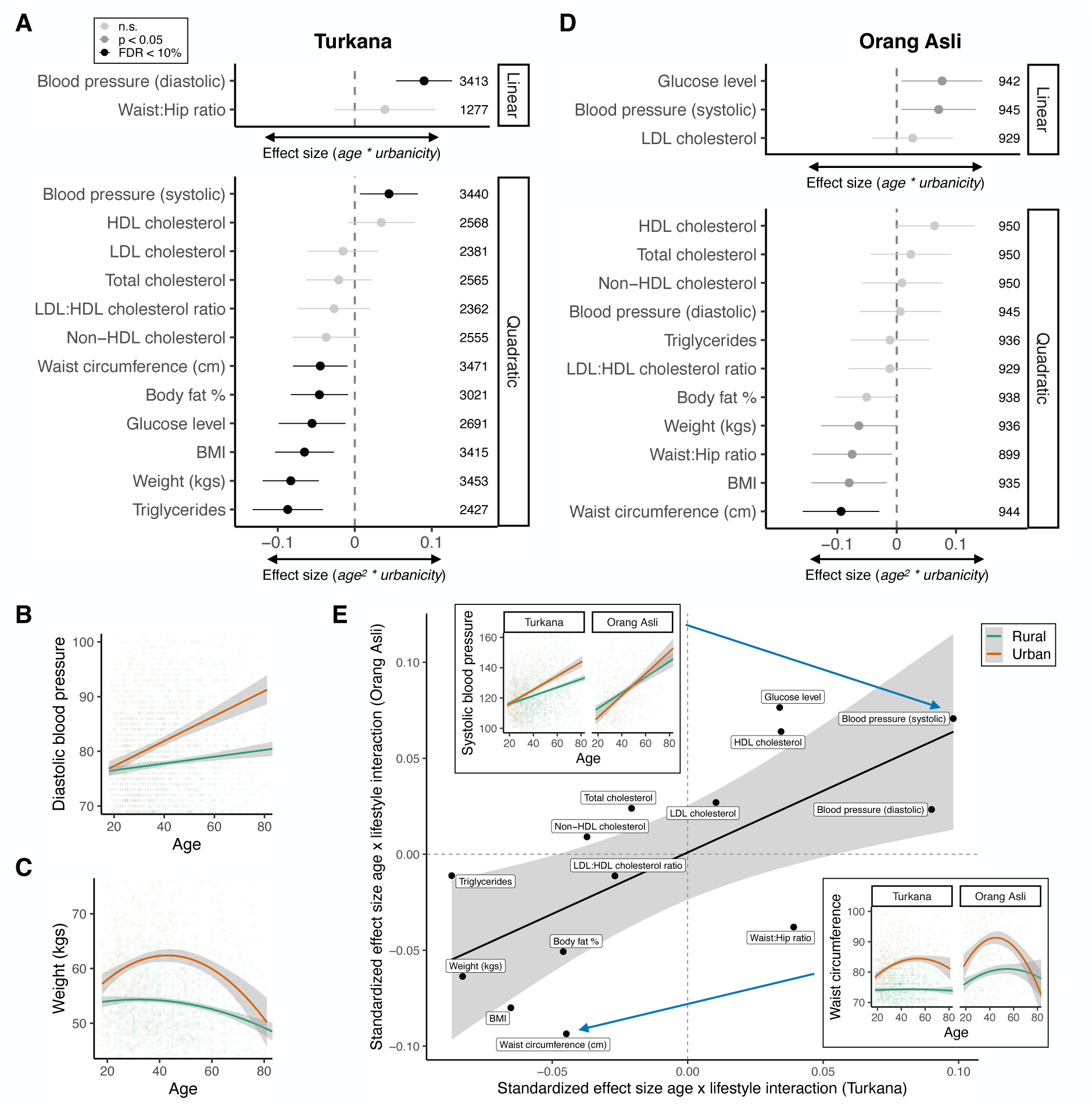
Lifestyle consistently exacerbates age effects across cardiometabolic biomarkers. Standardized effect sizes of interactive effects in (A) Turkana and (D) Orang Asli. Age x lifestyle effects shown for traits best predicted by a linear age term; age^2^ x lifestyle effects shown for traits best predicted by a quadratic age term. Colors show the level at which the effect is significant (shown in plot legend). Results from categorical outcomes (hypertension, overweight/obese) are not shown but are reported in Tables S2-3. Points further from the x = 0 line indicate greater lifestyle moderation across aging. Representative plot of age x lifestyle effects for (B) linear and (C) quadratic effects in Turkana. (E) Correlation of age x lifestyle in Turkana and Orang Asli. Age^2^ x lifestyle effects are plotted for biomarkers for which quadratic age terms were the best fit in both populations; linear age x lifestyle effects are plotted for all other biomarkers. Inset plots show examples of interactive effects for each population. We adjusted the axis limits in example plots to make age-related curves more easily observed, but all data points were still included in the analysis.

### Lifestyle-dependent age effects vary between sexes

In many countries, cardiometabolic health during aging diverges in sex-dependent ways. (60–62). As such, we sought to understand whether the age x lifestyle effects we observed differed substantially between females and males. Here, we focused on Turkana because our sample size was much larger, though we did recapitulate this analysis for Orang Asli (Turkana: Figure 3A-B, Table S5; Orang Asli: Figure S8, Table S6). We observed sex-dependent effects (where effects passed multiple hypothesis testing correction in one sex but had no nominal evidence in the other) for five biomarkers: body fat percentage, BMI, waist circumference, systolic blood pressure, and blood glucose. Concordant with prior research, among the 5 biomarkers with evidence for sex-dependent effects, females showed stronger age x lifestyle effects for those traits related to body composition (body fat percentage, BMI, and waist circumference), while males showed stronger age x lifestyle effects for blood glucose and systolic blood pressure.

**Figure 3:**
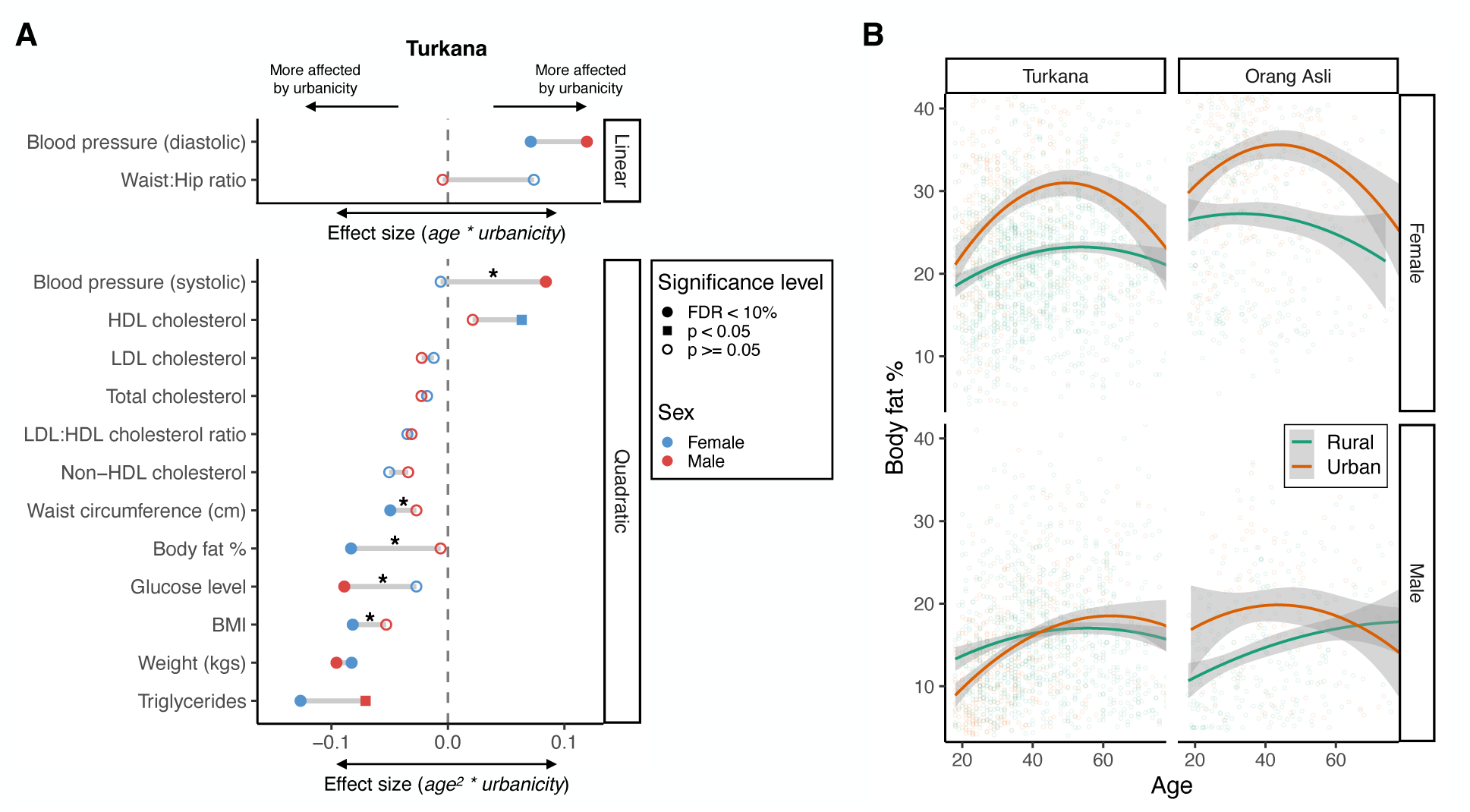
Sex differences in age x lifestyle interactions. (A) Interactive age x lifestyle (or age^2^ x lifestyle) effects from models of females (blue) or males (red) in Turkana. Shape of the dumbbell end points correspond to the significance level of the effect (legend on plot). Points further from the x = 0 line indicate greater lifestyle moderation of age effects. Asterisks denote significant differences in age x urbanicity effects between females and males (i.e., sex-dependent effects). Results from categorical outcomes are not shown but are reported in Tables S5-6. (B) Representative plot of age x lifestyle effects in female and male Turkana and Orang Asli.

### Similarity of age effects on cardiometabolic health in urban Turkana, Orang Asli, and US individuals

Following our observations that urbanicity amplified cardiometabolic age effects, we sought to understand whether these age x lifestyle effects reached magnitudes observed in a fully urban, industrialized setting. To test this, we again focused on our larger Turkana dataset (but see Figure S9 and Table S7 for recapitulation of general trends in the Orang Asli). We first binned Turkana into “urban” and “rural” following a natural break in the bimodal distribution of our continuous lifestyle metric (Figure 1B, urbanicity index = 27), and compared age effects in both Turkana groups to 10 random subsamples of US individuals from the National Health and Nutrition Examination Survey matched to the Turkana sample size and age distribution (Figure S10). When we compared age effects in rural Turkana to the US, we found differences in age slopes at 10/15 cardiometabolic biomarkers (FDR < 10% in ≥ 5 iterations); this included five biomarkers that were not significant in the within-Turkana age x lifestyle analyses (Figures 4A-C, S4; Table S8), but for which interaction effects were revealed by comparing to a fully urban, post-industrial dataset. Second, when we compared age effects in urban Turkana to the US, we surprisingly found differences for only diastolic blood pressure and hypertension (Figure 4A, S4; Table S8). This suggests that the urban environments Turkana experience are sufficient to generate health declines across age at largely similar magnitudes as those observed in a high-income, post-industrial country. Importantly, we note that while age effects may be similar in magnitude in urban Turkana and the US, the mean levels of most of these health biomarkers still differ dramatically, with much elevated risk factors among people living in the US (FDR < 10% for 14/15 tested biomarkers).

**Figure 4:**
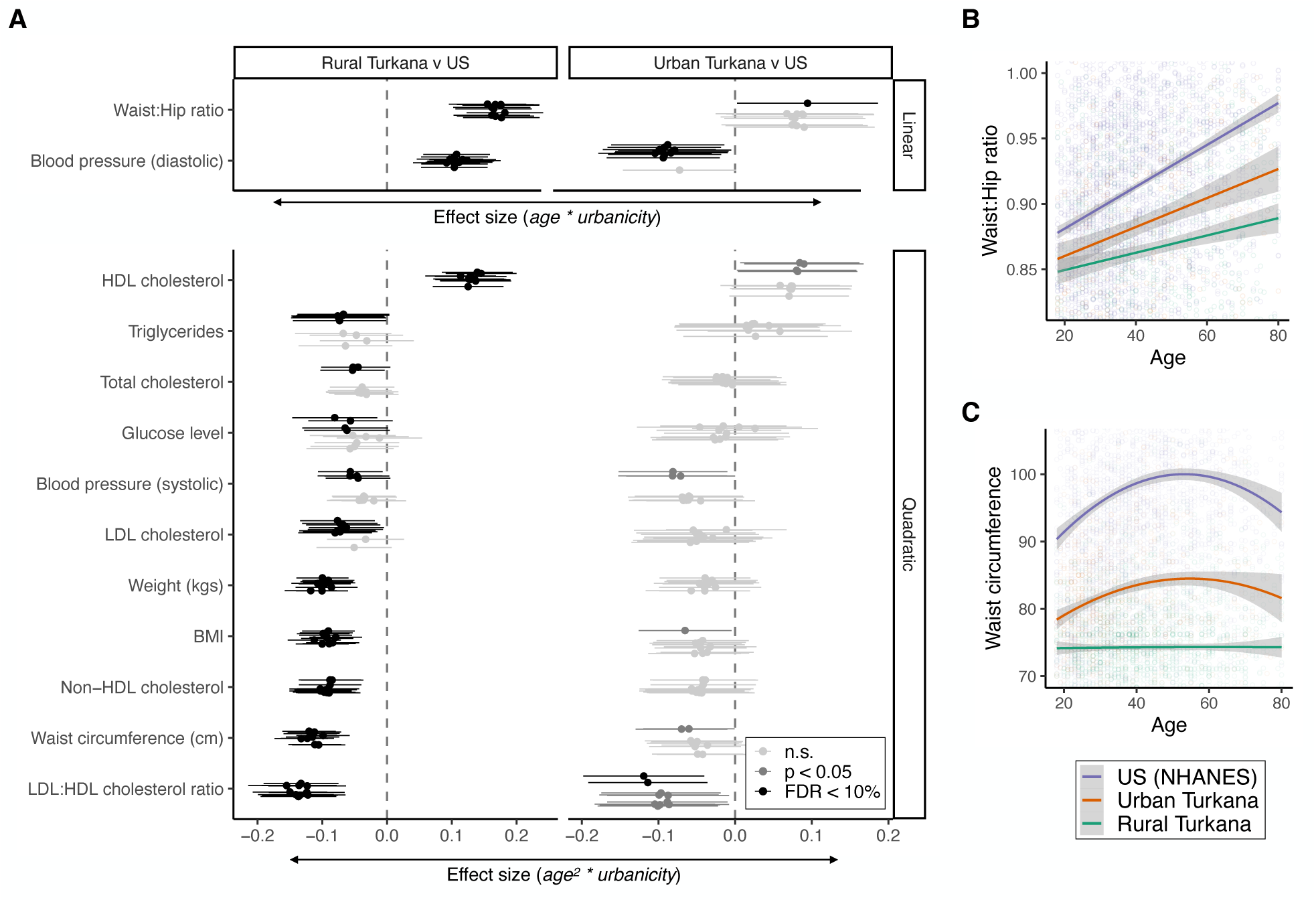
Similarity of age effects on cardiometabolic health in urban Turkana, Orang Asli, and US individuals. (A) Standardized effect sizes of age x lifestyle (linear age effects) or age^2^ x lifestyle (quadratic age effects) for each of the ten permutations of NHANES (US) data. The term used to model age was inherited from within-Turkana analyses. Color denotes the significance level each effect reached and is shown in the bottom right of plot A. Example plots of (B) age x lifestyle (C) and age^2^ x lifestyle. Purple shows US (NHANES) data. Turkana are characterized as urban (orange) or rural (green) following a natural break in the middle of the continuous urbanicity score (Figure 1B).

### Lifestyle effects on physiological and molecular biomarkers of aging vary between Turkana and Orang Asli

Given how strongly urbanicity perturbed age effects among different cardiometabolic health biomarkers, we wanted to understand whether urbanicity impacted aging processes at other biological levels. Investigations of lifestyle effects on epidemiological patterns rarely assess whether these effects extend to established biological mechanisms of aging. To address this, we first tested the extent to which grip strength and walking speed—markers of physiological function and frailty and consistent predictors of age-related morbidity and mortality (47, 63–65)—exhibited age, urbanicity, and age x urbanicity effects. We tested these effects on grip strength in each population and on walking speed in Orang Asli. As expected, these measures of physical function were correlated in Orang Asli (Pearson’s r = –0.33, p = 5.37 x 10^−16^). Consistent with previous work in both non-industrial and industrialized contexts (47, 63, 66, 67), we found that older individuals in both populations had weaker grip strength (Turkana: n = 673, age: β = –0.28, p = 9.42 x 10^−18^; Orang Asli n = 819, age: β = –0.14, p = 3.41 x 10^−7^; Figure 5A). However, we found mixed evidence that lifestyle impacts grip strength: in Turkana, individuals living in more urban environments had weaker grip strength (β = –0.2, p = 3.32 x 10^−11^) but among Orang Asli we did not find lifestyle effects. We also did not observe that age effects in either population were lifestyle dependent (p > 0.05; Figure 5A). Consistent with the grip strength patterns in Orang Asli, we observed that older Orang Asli walked more slowly than younger individuals (Orang Asli n = 661, age: β = 0.35, p = 1.66 x 10^−18^) and we did not observe main effects of lifestyle or age x lifestyle effects on walking speed. Together, these results suggest that the generalized physiological decline may not be accelerated by urban environments in the way we observe for cardiometabolic traits.

**Figure 5:**
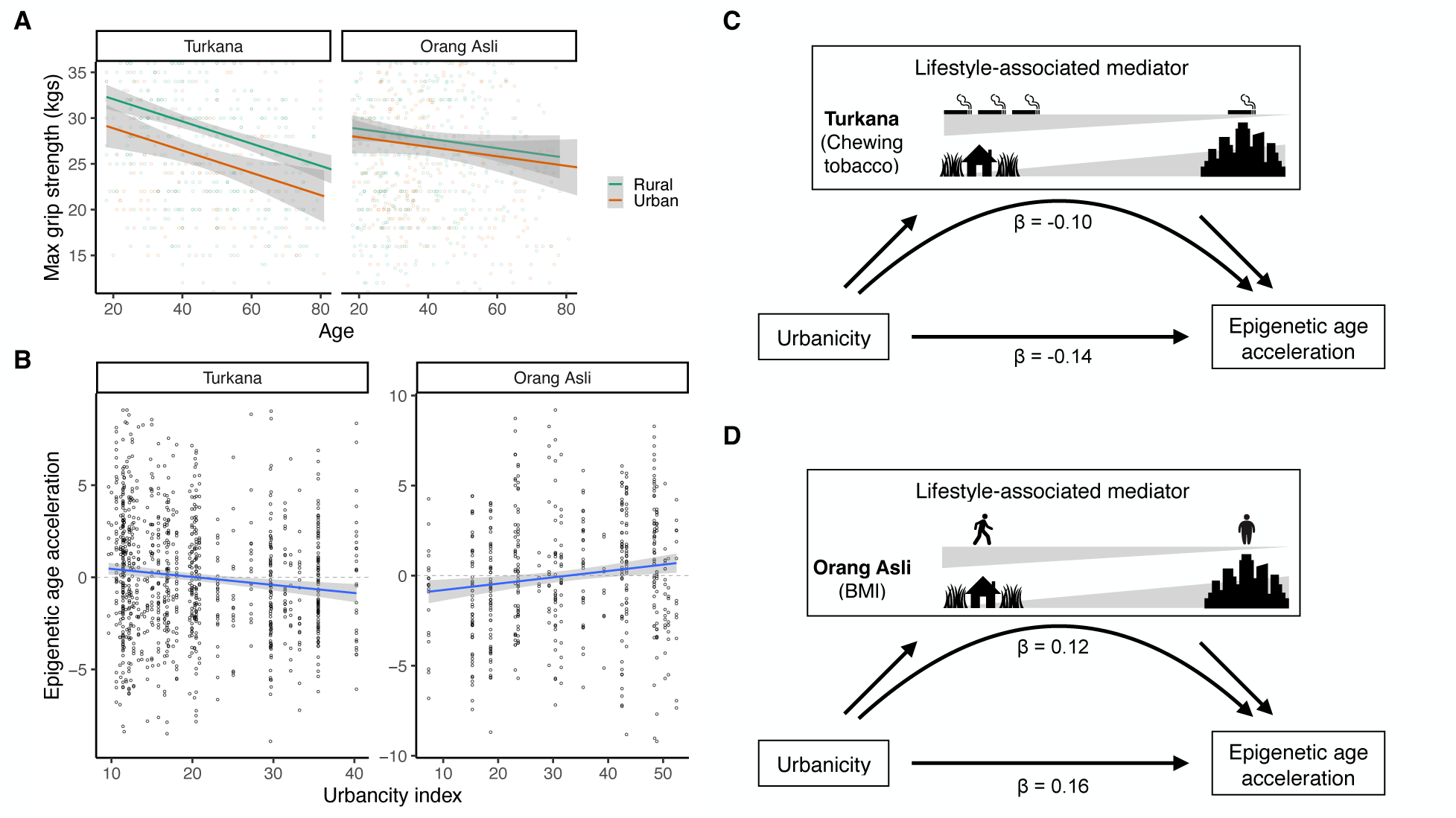
Lifestyle effects on cardiometabolic, molecular, and physiological aging are uncoupled in Turkana and Orang Asli. (A) Maximum grip strength declines across age differently in Turkana and Orang Asli, and this decline is not moderated by lifestyle in either group. (B) Urbanicity levels differently impact residual epigenetic age (population mean-centered epigenetic age acceleration plotted on y-axis), but these effects are partially mediated by distinct lifestyle-associated factors that proximately affect DNA methylation, including tobacco use in the Turkana and BMI–a proxy for diet and physical activity–in the Orang Asli. Schematic of mediators of the relationship between urbanicity and epigenetic age acceleration in (C) Turkana and (D) Orang Asli.

Next, we tested whether urbanicity accelerated biological aging using epigenetic clocks, which are known to reliably track age, predict age-related morbidities and mortality, and respond to lifestyle interventions (51, 68, 69). To do so, we generated genome-wide DNA methylation data for 1056 Turkana and 513 Orang Asli and, for each population, asked whether urbanicity was associated with epigenetic age acceleration, controlling for chronological age, sex, and cell type proportions. We focused on a clock we trained in our data (hereafter referred to as the ‘joint-population clock’) given evidence that existing clocks are less accurate when applied to populations whose ancestry differs substantially from the training dataset (4, 70, 71) (but see SI for information about other clock algorithms). Within Turkana, individuals whose predicted epigenetic age was older than their chronological age (i.e., age acceleration) tended to live in more rural, subsistence-based environments (β = –0.14, p_BH_ = 1.03 x 10^−4^) (Figures 5A, S11; Table S9). However, in Orang Asli, epigenetic age acceleration followed the opposite trend, and was positively associated with urbanicity (β = 0.16, p_BH_ = 5.64 x 10^−3^) (Figures 5B, S11; Table S9).

We were surprised that urbanicity had seemingly opposing effects on epigenetic age acceleration in Turkana and Orang Asli (Figure 5B), but we noticed that the differences in the direction of urbanicity on epigenetic aging between the two populations were driven by rural individuals (Figure S12). Given that epigenetic clocks are sensitive to lifestyle factors that might have different associations with urbanicity in each population, we tested for mediation for BMI and tobacco use, which have been repeatedly associated with epigenetic age acceleration and which covary with lifestyle in distinct ways in each group (Figure 5C-D) (72–75). In Turkana, rural individuals use tobacco at a 1.13x higher rate than urban individuals, while in Orang Asli, tobacco use is similar across the urbanicity gradient (urban = 28%, rural = 30%). We found that this variation in tobacco use mattered for biological aging, mediating 23.3% of the effect of urbanicity on epigenetic age (decreased effect of urbanicity: β = –0.11, p = 1.39 x 10^−3^; boot-strapped CIs: –0.058, –0.008) (Figure 5C). In Orang Asli, urban individuals exhibit 18.5% higher average BMI than rural individuals, an effect that is much more subtle in Turkana (where urban individuals exhibit 4.19% higher BMI than rural individuals). As expected, we found that this variation in body composition mattered in Orang Asli: BMI mediated 26.0% of the relationship between urbanicity and epigenetic age acceleration (adjusted effect of urbanicity: β = 0.12, p = 1.88 x 10^−2^; boot-strapped CIs: 0.01, 0.08; Figure 5D). These results emphasize that the details of lifestyle transitions can be heterogeneous despite broad epidemiological similarities, with consequences for key processes at the biological level.

## Discussion

We set out to answer two main questions. First, to what extent is cardiometabolic morbidity in old age an unavoidable consequence of physiological aging and long human lifespans or, alternatively, a result of a lifetime of exposure to modern, urbanized environments that are prone to higher cardiometabolic risk (23, 76)? Second, if urban exposures produce or exacerbate cardiometabolic age effects, does urbanicity also impact aging processes throughout the body, or instead, are these effects limited to specific physiological systems (i.e., cardiometabolic)? Working with two long-term studies, we find that age effects on cardiometabolic health are highly modifiable by lifestyle, and largely “generated” by urban lifestyles within both groups. However, these patterns do not consistently extend to biomarkers of biological age: grip strength and walking speed exhibited no evidence of lifestyle-dependent age effects, and epigenetic age was oppositely accelerated by Orang Asli urban environments and Turkana rural environments. The consistency of age x lifestyle effects on cardiometabolic health biomarkers we observed across two groups living in very different ecologies and socio-political settings (45) underscores that many transitioning populations are likely at risk for similar cardiometabolic effects (33, 34, 42, 77, 78) even though urbanicity’s effects on biological aging may be more population-specific (79).

While recent studies have suggested strong effects of age on many NCDs (1), but in contexts where individuals have experienced urban environments since early age, age is essentially fully correlated with environment. Our powerful within-group framework reveals that much of the effect that has previously been attributed to age may instead be an effect of lifestyle, consistent with previous work focusing on non-industrial populations alone (25, 31–33). Importantly, our work in this area includes a pastoralist population, in addition to horticulturist and hunter-gatherer populations, which are more well-represented in the health literature (25, 27, 28, 43, 64) ((29, 80–82) for studies of cardiometabolic health in pastoralists).

While urbanization consistently amplified age-related epidemiological patterns, its effects on epigenetic aging diverged notably: urbanization was associated with accelerated biological age in Orang Asli, but with deceleration among the Turkana. Rather than dismissing this as technical noise, we suggest this divergence could actually be meaningful. It highlights that biomarkers of biological aging—here, DNA methylation clocks—may be sensitive not only to age or pathology, but to specific environmental, cultural, or developmental exposures that differ between groups. This suggests that molecular and physiological aging may be at least partially uncoupled from epidemiological risk of aging, such that cardiometabolic decline can sometimes occur without corresponding acceleration in epigenetic age. Our investigation also revealed substantial heterogeneity in epigenetic aging that was unexplained by urbanicity or the environmental mediators we tested and we advocate for more granular analyses of how DNA methylation is affected by urbanicity. In particular, analyses at the CpG-level may be more well-suited to disentangling causal directions, as a major caveat of epigenetic clock analyses is ambiguity in the degree to which these measures capture causes or consequences of aging (83).

We observed few age by lifestyle interactions when comparing Turkana living in urban areas and US individuals, suggesting that the extent of urbanicity experienced by Turkana living in urban areas is already enough to generate similar age-effects on cardiometabolic health as a fully industrialized setting. However, the average values for most of these traits are still dramatically different between the two populations. Notably, both the intercept and slope–which are strongly affected by urbanicity–of age effects practically impact whether an individual will reach disease thresholds for cardiovascular diseases. Given the pace of urbanization and industrialization around the world, analyses like these are imperative for understanding how quickly lifestyle transitions can contribute to disease burden.

We acknowledge several limitations and areas for future work. First, using BMI as a cardiometabolic outcome is imperfect as BMI is inherently biased with respect to height and there is mixed evidence about whether BMI under 30 has deleterious effects during aging (84, 85). However, we include it here because it has been used in many previous studies. Future work focusing on additional fine-grained measures of metabolic function (e.g., lipidomics) or clinical measures (e.g., coronary artery calcification) could be especially fruitful. A second limitation is the potential for survivorship bias, such that healthier individuals are more likely to persist into older age. We do not expect that survivorship bias drives the minimal age-associated differences we observe in rural individuals, given the ranges of cardiometabolic health traits we observed. Survivorship bias has also been put forward as a reason explaining the quadratic age-health biomarker trajectories, but longitudinal studies have reinforced quadratic age effects, especially on body composition-related traits, within individuals (12). Longitudinal data collection in transitioning populations is greatly needed to add to this body of literature, and to generally identify the causes and consequences of diverse exposomes on healthy aging. Lastly, we trained our epigenetic clock to predict chronological age, rather than morbidities or mortality or aging trajectories. “Second”-or “third-generation” clocks trained on health biomarkers, mortality, or the pace of aging, respectively, can be more predictive of disease development or biological processes that contribute to death, and also tend to be more sensitive to pertinent social and behavioral variables (69, 86, 87). We look forward to the future development of second and third generation clocks that incorporate environmentally and genetically diverse individuals in the training set and will thus be well-suited to study lifestyle effects on biological aging in populations around the globe.

Taken together, our results demonstrate that urban environments magnify or generate age-associated differences in diverse cardiometabolic biomarkers including blood lipids, body composition, and blood pressure, yet this environmental amplification of age effects does not occur uniformly across other age-associated biological traits. To some degree, our results call into question the Geroscience Hypothesis, which posits that slowing the pace of biological aging can mitigate diseases of aging and prolong healthspan (16), and suggest that the effects of lifestyle may be partially decoupled across biological systems. Additional investigations are needed to understand how the timing, duration, and magnitude of environmental exposures impact the extent to which age-effects are observed across multiple biological levels. Indeed, the trajectories of individuals’ cardiometabolic, musculoskeletal, physiological, and molecular traits throughout aging (88), and the extent to which they co-vary, may emerge as key indicators of vulnerability and functional decline.

## Methods

### Turkana, Orang Asli, and data collection

The Turkana people have inhabited Turkana County in Northwest Kenya from the early 18th century and are traditionally nomadic pastoralists who currently form the second largest pastoralist group in Kenya. The diet of pastoral Turkana is derived primarily (∼70-80%) from milk and other animal products (42, 89). Turkana County is an exceptionally arid area, receiving an average of <400 mm of rainfall per year (90). Over the last several decades, Turkana County and much of northern and western Kenya have seen rapid, widespread infrastructure development. This has resulted in the growth of several urban centers near or in traditional Turkana lands, the growth of small-scale market trade throughout the region, and many economic, social, and cultural changes for the Turkana. Increased infrastructure and economic opportunities in the region have seen many Turkana move to urban centers within northern and central Kenya, or remain in rural areas but increasingly participate in the market economy (42, 53).

We used data collected by the Turkana Health and Genomics Project (THGP) (March 2018-November 2023) described in (42). Following meetings with community leaders and members, the THGP project recruited consenting, healthy adults (>18 years) to participate in structured surveys with a research team member who was familiar with the local community and language of the participant (e.g., Turkana, Swahili, English). All community members, regardless of whether they participated in the project, were provided free healthcare if desired. Individuals who wished to participate took part in structured interviews asking about demography, subsistence and labor practices, material wealth and housing infrastructure, diet, health, early life experiences, and relocation history. Written, informed consent was obtained from all participants after the study goals, sampling procedures, and potential risks were explained to participants in their native language by researchers. Participants were also invited to participate in the measurement of 14 cardiometabolic phenotypes, including waist circumference, body fat percentage, weight, BMI, waist-to-hip ratio, total cholesterol, high-density and low-density lipoprotein (HDL, LDL) cholesterol, non-HDL cholesterol, LDL-to-HDL ratio, triglycerides (mg dL), glucose level (mg/dL), and systolic and diastolic blood pressure (mm Hg). Data collection is further described in (42) and (45). Demographic summaries of participants are included in Figure 1A and Table S1. The THGP was approved by the Princeton University Institutional Review Board for Human Subjects Research (Institutional Review Board no. 10237) and Maseno University (approval number MUERC-00519-18).

“Orang Asli” refers broadly to the Indigenous peoples of Peninsular Malaysia. The Orang Asli are typically categorized into three broad groups (the Negrito [Semang], Senoi, and Aboriginal Malay) and 19 culturally distinct ethnolinguistic groups, and while these three subgroups are genetically distinct, the Orang Asli are generally more genetically similar to each other than to ethnic Malays or other Asian populations (91) (see Table S1 for details of ethnolinguistic groups). Collectively, the Orang Asli traditionally practiced varied subsistence histories consisting of foraging, swidden agriculture, trade of rainforest products, or a combination thereof, and these practices were ubiquitous among Orang Asli until the 1950’s and remain common in some areas today (54, 56, 92). Over the last several decades, Malaysia has undergone one of the fastest rates of urbanization, deforestation, and industrialization worldwide (92), which, combined with government efforts to assimilate the Orang Asli into the national economy, has resulted in Orang Asli experiencing a wide gradient of lifestyle difference from individuals living in remote interior rainforest villages and practicing small-scale, subsistence level practices such as foraging and hunting to Orang Asli villages being surrounded by urban or peri-urban development and individuals participating in the market economy instead of, or in addition to, traditional practices (43, 56).

We used data collected by the Orang Asli Health and Lifeways Project (OA HeLP) that were collected between March 2020 and October 2024. OA HeLP works in Orang Asli communities and performs similar sampling and data collection as the THGP and also provides free healthcare to Orang Asli and community members regardless of participation status. Sampling procedures for OA HeLP are further described in (43) and (45) and are briefly described here. After consent from community leaders and hosting community-wide informational sessions, OA HeLP recruited adults to participate in similar interviews. The study aims, procedures, and potential risks were explained to all participants and written, informed consent was obtained from all participants. The OA HeLP was approved by the Medical Review and Ethics Committee of the Malaysian Ministry of Health (protocol ID: NMRR-20-2214-55565), the Malaysian Department of Orang Asli Development (permit ID: JAKOA.PP.30.052 JLD 21) and the Institutional Review Board of Vanderbilt University (protocol ID: 212175).

### Measuring cardiometabolic biomarkers

Participant interviews and biometric data collection is performed similarly by the THGP and OA HeLP. During participant interviews, multiple cardiometabolic and biometric phenotypes were measured, including standard anthropometric measurements such as waist circumference, hip circumference, body fat percentage, weight, standing height, systolic and diastolic blood pressure, and a panel of blood lipids. Hip and waist circumference were measured three times and averaged to generate a final value. Body mass index (BMI) was calculated by dividing body weight (kgs) by height (m^2^) and participants were categorized as overweight/obese following the standard cut-off of BMI ≥ 25 (93). Body fat percentage was measured using the Omron HBF-306C Handheld Body Fat Loss Monitor in Turkana and a TANITA BC-558 FDA Cleared Ironman Segmental Body Composition Monitor was used in Orang Asli. Blood pressure was measured with the Omron 10 Series Wireless Upper Arm Blood Pressure Monitor following standard manufacturer procedures in Turkana, and the Omron 5 Series Upper Arm Blood Pressure Monitor was used for the Orang Asli. Participants in both populations were characterized as hypertensive if systolic blood pressure was greater than 135 and diastolic blood pressure was greater than 85. To reduce the possible effects of ‘white coat syndrome’, blood pressure was measured twice in both populations and we used the second measure in our analyses. The Jamar Hydraulic Hand Dynamometer was used to measure grip strength separately for the left and right hands. Walking speed was measured as the number of seconds that participants took to walk 10 meters at their maximum speed. Individuals who required mobility assistance devices were excluded from the collection walking speed data. During sampling, participants were invited to provide a small (approximately 15mL) venous blood sample. One mL of this sample was used to measure lipids including total cholesterol, triglycerides, and high– and low-density lipoproteins via the CardioCheckPlus panel from PTS Diagnostics. Non-HDL cholesterol was quantified by subtracting HDL from total cholesterol and LDL was calculated using the Friedewald Equation (LDL cholesterol = total cholesterol – HDL cholesterol – triglycerides/5). As the CardioCheckPlus panel does not reliably detect triglycerides or total cholesterol under 50 mg/dL, we removed values under 50 from analysis. Blood glucose was measured from a drop of blood via finger prick using the OneTouch system. Further methods can be found in (42, 43, 45).

### Data filtering

Individuals were excluded from analyses if they met any of the following criteria: (i) were missing age or sex information or (ii) were currently pregnant, and for Turkana if they (iii) were taking medications for hypertension or diabetes (n = 16). We removed individuals from the THGP dataset if they were of non-Turkana tribe affiliation. We also removed clearly erroneous values (e.g., missing decimal places). Next, we allowed a large (n = 5965), publicly available and nationally representative dataset of US individuals–the National Health and Nutrition Examination Survey (NHANES) database from the Center for Disease Control (46)–to inform the universe of possible lower bounds for all continuous biomarkers. While biomarker values from Turkana and Orang Asli generally fell between this lower bound and the maximum represented in NHANES, we removed values lower than the NHANES lower bound. We allowed values to surpass the maximum in NHANES for most measures if they were biologically possible. However, we limited waist-to-hip ratio values to be within the range observed in NHANES. Ranges of all continuous biomarkers per population are reported in Table S10 and the distributions of all biomarker values for Turkana and Orang Asli are in Figures S2 and S3, respectively. Before statistical analyses, all continuous biomarkers were mean centered and scaled by standard deviation. Consequently, all reported effect sizes are standardized and represent the effect of a given variable on the outcome in terms of increases in standard deviations.

### Urbanicity scale generation

As Turkana and Orang Asli span wide lifestyle gradients from traditional subsistence-level strategies to living in highly urban city centers, we generated a continuous measure of urbanicity per sampling location. Measuring urbanicity at the location-level is advantageous because it allows for individuals who may be missing individual-level data to be included in analysis and it provides a representative depiction of the resources available within a community (e.g., individuals may not own a mobile phone but still have access via family or friends). We quantify urbanicity using a scale that was first proposed by (44) and measures features of the built environment, infrastructure, and population density at the location-level. This scale was tested in Turkana and Orang Asli (45) and found to predict cardiometabolic health better than other continuous measures of urbanicity. In our analyses, this scale ranges from 9.5 to 40.26 in Turkana and 4.11 to 42.45 in Orang Asli. We observed a bimodal distribution in urbanicity in both populations with a break at approximately 27 (Figure 1B). We use this cut-off of 27 for analyses that required a binary rural/urban designation and for plotting purposes to show age x lifestyle interactions.

### Within-population age x lifestyle effects

First, we investigated the extent to which lifestyle affected the effects of age (i.e., age-associated differences), as well as mean effects of age and lifestyle, within the Turkana and Orang Asli separately. To do so, we modeled each of our 16 cardiometabolic outcomes using the four models detailed in Figure 1C, and controlling for sex. These models tested additive effects of age and lifestyle and interactive age x lifestyle effects using both linear and quadratic effects of age, as many of the traits we investigated have been found to change in non-linear patterns across age (12). We modeled continuous biomarkers using linear models and the *lm* function in R and categorical variables (hypertension and overweight/obese) using a generalized linear modeling framework with the *glm* function and binomial error distribution and logit link function (94). After modeling each trait with these four models, we quantified the model that best described age and lifestyle patterns for each cardiometabolic trait using a model selection approach by comparing model fit (AIC) of the four models. We performed this model selection in a hierarchical manner by first asking whether models of the most complexity (quadratic interactive) showed a significant effect of age^2^ x lifestyle (FDR < 10%) and a ΔAIC > 2 between this model and the next most complex (linear interactive). If either criteria was not met, we discarded that model as the best fit. We used an FDR threshold of 10% because this would show a trend towards an interactive effect that might be missed with a more stringent threshold. We performed this test iteratively by model complexity, necessitating that the following effects were significant (FDR < 10%) per model type: quadratic interactive (age^2^ by lifestyle; most complex), linear interactive (age by lifestyle), quadratic additive (age^2^), linear interactive (default; least complex). For all analyses, we corrected for multiple hypothesis testing using a Benjamini-Hochberg FDR from the *p-adjust* function in R (94). In Turkana, our model selection approach revealed that three biomarkers were best predicted by a linear age term: waist-to-hip ratio, diastolic blood pressure, and hypertension, while the rest were best predicted by a quadratic age term. In Orang Asli, LDL cholesterol, systolic blood pressure, and blood glucose were best predicted by a linear age term and the rest by a quadratic age term. In later analyses, we used linear or quadratic terms to model age x lifestyle interactions and these were inherited from the model of best fit for the within-population analyses. All statistical analyses were done using R version 4.42 (94).

### Age effects in Turkana practicing traditional pastoralism

To understand which cardiometabolic phenotypes exhibit age-related differences in individuals who are not, or minimally integrated into the market-economy, we investigated age effects among Turkana who practice traditional pastoralism. We focused this investigation in Turkana because we could identify individuals living subsistence-based, pastoralist lifestyles with minimal to no market-integration. Controlling for sex, we asked if there were age effects for 15 biomarkers within a subset of 330 individuals who (i) practice and rely on subsistence-level livestock herding, (ii) live in rural areas in or around Turkana County, and (iii) do not or rarely consume sugar, salt, or oil (criteria adapted from (42)). We did not test waist-to-hip ratio as we did not have hip circumference measurements for this subset of individuals. As expected, the traditional pastoralists lived in places that had extremely low urbanicity (range: 10-15.4, i.e., the lowest 13% percentile of urbanicity observed among all Turkana in our dataset). As this subset of individuals showed extremely low variation in urbanicity, we tested only for age effects and not age x urbanicity effects. Additionally, we did not necessarily expect that age patterns among traditional pastoralists would be the same as those for Turkana living in urban areas or partially integrated in the market economy (i.e., peri-urban). Within this subset of individuals, we compared models with age as a linear and quadratic effect and report age effects from the model that best fit the data (ΔAIC < 2; Figure S7, Table S4).

### Sex differences in age x lifestyle effects

To test whether females and males differed in the extent to which urbanicity impacts age-related cardiometabolic health, we modeled age x lifestyle effects within females and males separately in each population using the age term from the best fit model from our within-population model selection approach. We then identified biomarkers for which the age x lifestyle effect was significant (FDR < 10%) in one sex but showed no detectable evidence in the other sex (p ≥ 0.05) or for which the sexes showed differences in the direction of effect (FDR < 10%), indicating a sex-dependent effect (95, 96). We acknowledge that biological sex comprises a suite of biological attributes and that binary assignment and analysis may not reflect the full diversity of this complexity. Additionally, while there is some malleability in concepts surrounding sex and gender in Turkana and Orang Asli culture, gender presentation and roles among Turkana and Orang Asli are strongly stratified by biological sex.

### Comparing age x lifestyle effects between Turkana and Orang Asli and a high-income, post-industrialized population

To compare Turkana and Orang Asli to a high-income, post-industrialized population, we used publicly available data from the US NHANES database (46). The data we used were collected between 2017-2020. We matched survey and biomarker variables between the US and Turkana datasets as closely as possible. Variables were pulled from the following datafiles: P_DEMO.XPT, P_BPXO.XPT, P_BPQ.XPT, P_BMX.XPT, P_HDL.XPT, P_TRIGLY.XPT, P_TCHOL.XPT, P_CBC.XPT, P_DIQ.XPT, P_GLU.XPT, P_SMQ.XPT, P_SMQRTU.XPT, and P_UCPREG.XPT. We removed individuals who were younger than 18 years, were pregnant, or were taking medications for cholesterol, diabetes, or blood pressure management. Additionally, NHANES includes three measures each of systolic and diastolic blood pressure, and–to remove likely errors–we first removed values that differed by more than 30% of the mean of the other two values, and we then generated a mean measurement for each blood pressure trait. We characterized overweight/obese and hypertension as binary variables with the same method as for Turkana. We excluded US individuals on medications to manage cholesterol, blood pressure, and diabetes, likely biasing our American sample to healthier than average individuals and producing conservative estimates in the extent to which post-industrial environments affect cardiometabolic biomarkers. As the NHANES dataset was larger than and had a slightly different age distribution than the Turkana, we generated age-matched datasets to match the age distribution and sample size of the Turkana dataset. We separated age into 13 bins of even sizes for ages 20-80 and one bin of 18-20 years. We performed random subsampling of each NHANES age bin to match the number of Turkana for that age range. Eighty was the maximum age in the NHANES dataset and as such, we removed individuals greater than 80 years old from the Turkana data for all comparative analyses between the two populations (n = 3693) and used this sample size for NHANES subsampling. As random sampling can inherently create unintentional biases in or variation in results, we performed ten permutations and used these ten permutations in all downstream analyses.

We used these US data to investigate age x lifestyle effects between rural and urban Turkana (or Orang Asli) by modeling each of the 15 cardiometabolic traits (NHANES does not include body fat percentage) by age (or age^2^), lifestyle, age x lifestyle (or age^2^ x lifestyle), and sex. Specifically, we asked whether the slopes of age effects were different between the rural/urban Turkana/Orang Asli and the US. If the US showed a slope more extreme than that in urban Turkana/Orang Asli, this would suggest that increasing levels of urbanicity further affect cardiometabolic health across aging. We observed that several traits trended towards having significantly different slopes between the urban individuals and US, and we next tested whether rural individuals showed different age slopes than the US. In cases where rural, but not urban, individuals showed differences in age effects from the US, this would suggest that urbanicity affects cardiometabolic aging but that these effects are only observed at certain ranges of the urbanicity gradient. We tested for age x lifestyle effects with the age term from our within-population model selection approach. We performed multiple hypothesis test corrections on all models.

### Age and lifestyle effects on grip strength and walking speed

Finally, we sought to quantify whether two measures of muscle strength and frailty varied with age and urbanicity and whether urbanicity moderated age effects on grip strength in both populations (n Turkana = 673, n Orang Asli = 819) and walking speed in Orang Asli, for which we had sufficient data (n = 663). We did so by quantifying the extent to which grip strength and walking speed varied across age. We quantified grip strength as the maximum value an individual obtained between the left and right hand. Walking speed was quantified as the number of seconds that an individual took to walk 10 meters at their maximum speed. We first asked whether there were main effects of age and urbanicity in both populations, modeling grip strength separately in each population, with age and urbanicity as additive terms in our model. We then asked whether age-related differences in strength were moderated by lifestyle with an age x urbanicity effect. We modeled age as a linear term for all models.

### Lifestyle effects on epigenetic age acceleration

We quantified DNA methylation (DNAm) via enzymatic methylation sequencing using a recently described target panel that profiles methylation at ∼4 million CpG sites (97). DNA was extracted from whole blood samples provided by THGP using the Zymo Research Quick-DNA miniprep kit and from peripheral blood mononuclear cells isolated from whole blood provided by OA HeLP using the Quick DNA 96 Kit from Zymo Research. DNA methylation library preparation was done using the NEBNext® Enzymatic Methyl-seq kit and were sequenced on the NovaSeq X at the Vanderbilt Technologies for Advanced Genomics (VANTAGE) Core. Library preparation and sequencing is detailed in (97). Sequencing data were processed using the Illumina DRAGEN Methylation Pipeline v4.1.23 and we extracted percent methylated values (methylated counts / total counts) for each CpG site per sample. We excluded samples with extremely low read count (<1000000), mapping percent < 70, or CHH methylation < 10% and we excluded sites with a median coverage of <5X in <75% of samples. We also limited this dataset to CpG sites within the target probes regions (+/-200 bp) and to CpGs within the Illumina EPIC array, as this array has been used to construct the majority of commonly used epigenetic clocks (n CpGs Turkana = 616422, Orang Asli = 652349) (49, 50, 86, 98). We also removed samples and sites with high missingness (> 95%).

We estimated proportions of major blood cell types using the EpiDish package in the R environment (99). Estimated values were correlated with values from complete blood counts, which we had for a subset of individuals in each population (Turkana lymphocytes: r = 0.58, p = 1.28 x 10^−81^, neutrophils: r = 0.58, p = 9.54 x 10 ^−81^; Orang Asli lymphocytes: r = 0.36, p = 6.4 x 10^−15^).

Epigenetic clocks are known to be sensitive to recent genetic ancestry because DNAm is partially controlled by genotype (4, 70, 100). As such, epigenetic clocks generally do not predict age as well on datasets with different recent genetic ancestries than those which they were trained on (4, 70, 71). Most commonly used epigenetic clocks were trained on data primarily from individuals of European ancestry (49, 50, 86, 101). Therefore, we trained an epigenetic clock on the DNAm data we generated from the Turkana and Orang Asli. To do so, we further filtered each dataset to remove CpGs associated with genetic ancestry (i.e., meQTLs) by removing CpGs within 50 base pairs of meQTL SNPs (SNPs associated with variable methylation) detected in data from the 1000 Genomes Project, resulting in 598078 CpGs for the Turkana dataset and 631837 for the Orang Asli dataset. We then merged these datasets, keeping only CpGs sites present in both datasets (n CpGs = 596462). We imputed any missing data and used the standard workflow from the PC-Clock R package to generate a PC-based epigenetic clock (i.e., the joint-population clock) and obtained epigenetic age acceleration values with the calcPCClocks_Accel function.

Despite the confound with genetic ancestry, we also predicted epigenetic age from 10 established and previously validated clocks to allow our results to be comparable to previously published work. Specifically, we predicted epigenetic age from the CpG-based Horvath, Hannum, Levine, and telomere length (TL) clocks, the Horvath, Hannum, TL, PhenoAge, and GrimAge PC-based clocks, and DunedinPACE using the methyclock, DunedinPACE, and PC-Clock R packages, respectively (86, 98, 102). Epigenetic age acceleration (i.e., residuals, hereafter EAA) was calculated for each clock from their respective R packages, except DunedinPACE which estimates pace of aging. As expected, we observed that the joint-population clock and PC-based clocks generally better predicted chronological age than the first-generation CpG-based clocks (correlations with chronological age shown in Figure S11A-B).

To test how urbanicity affects epigenetic age acceleration, we modeled EAA and DunedinPACE as a function of chronological age, sex, urbanicity, and estimated proportion of major blood cells. We controlled for estimated lymphocyte and neutrophil proportions in analyses of Turkana data and for estimated lymphocyte proportions in analyses of Orang Asli data. We modeled EAA separately per population for all clocks to more easily assess within-population trends. Following our observation that urbanicity was differently associated with EAA in each population, we sought to understand whether salient environmental variables mediated the effects we observed. We focused our mediation investigation on tobacco use in Turkana and BMI in Orang Asli because we knew that each variable was associated with urbanicity within each population and differed between the populations (i.e., rural Turkana use tobacco at higher frequencies than rural Orang Asli). To directly compare the results in each population, we tested each variable for mediation effects in both populations. To implement the mediation analyses, we first estimated the indirect effect of urbanicity on EAA through each mediator. We compared the effect size of urbanicity in these ‘adjusted’ models to ‘unadjusted’ models of the direct effect of urbanicity without the mediator (described above). If the variable is a strong mediator, then the effect of urbanicity will be reduced in the adjusted models because the variance attributed to urbanicity in the unadjusted model is explained by the mediating variable. To test the significance of each mediating variable within each population, we estimated the decrease in the effect size of urbanicity between the two models across 1000 bootstrap resampling iterations using the boot package in the R environment (103). We determined the variable to be significant if the confidence intervals did not overlap with 0. We also quantified the percent change in the urbanicity effect between the two models (i.e., the percent of the effect of urbanicity on EAA that was mediated by the mediating variable) using the effect size estimated from the full dataset without bootstrapping.

### Data Sharing

Our highest priority, and that of the THGP and OA HeLP, is to minimize risk to study participants. As such, both projects adhere to the “CARE Principles for Indigenous Data Governance” (Collective Benefit, Authority to Control, Responsibility, and Ethics) and are committed to the “FAIR Guiding Principles for scientific data management and stewardship” (Findable, Accessible, Interoperable, Reusable). To adhere to these principles while minimizing risks, individual-level data are stored in the protected data repositories and are available through restricted access. Requests for de-identified, individual-level data should take the form of an application that details the exact uses of the data and the research questions to be addressed, procedures that will be employed for data security and individual privacy, potential benefits to the study communities, and procedures for assessing and minimizing stigmatizing interpretations of the research results. Requests for de-identified, individual-level data will require institutional IRB approval (even if exempt). THGP and OA HeLP are committed to open science and the project leadership is available to assist interested investigators in preparing data access requests (see turkanahgp.com and orangaslihealth.org for further details and contact information). Data needed to evaluate the conclusions in the paper are present in the paper and/or the Supplementary Materials. Code used to perform analyses described herein is available on our GitHub (https://github.com/mwatowich/Cardio_ageXlifestyle) and code used to generate lifestyle scales is available at: https://github.com/mwatowich/Multi-population_lifestyle_scales.

## Data Availability

Our highest priority, and that of the THGP and OA HeLP, is to minimize risk to study participants. As such, both projects adhere to the "CARE Principles for Indigenous Data Governance" (Collective Benefit, Authority to Control, Responsibility, and Ethics) and are committed to the "FAIR Guiding Principles for scientific data management and stewardship" (Findable, Accessible, Interoperable, Reusable). To adhere to these principles while minimizing risks, individual-level data are stored in the protected data repositories and are available through restricted access. Requests for de-identified, individual-level data should take the form of an application that details the exact uses of the data and the research questions to be addressed, procedures that will be employed for data security and individual privacy, potential benefits to the study communities, and procedures for assessing and minimizing stigmatizing interpretations of the research results. Requests for de-identified, individual-level data will require institutional IRB approval (even if exempt). THGP and OA HeLP are committed to open science and the project leadership is available to assist interested investigators in preparing data access requests (see turkanahgp.com and orangaslihealth.org for further details and contact information). Data needed to evaluate the conclusions in the paper are present in the paper and/or the Supplementary Materials.

## Acknowledgments

First and foremost, we thank Turkana and Orang Asli community members and leaders for their contributions and support of our scientific work. We also thank all previous members of the Turkana Health and Genomics Project and the Orang Asli Health and Lifeways Project. We are also grateful to the staff of Mpala Research Centre and the Kenya Medical Research Institute for their essential support.

## Author contributions

M.M.W., Y.A.L.L., D.M., S.N.N., K.S.N., V.V.V., I.J.W., J.F.A., M.G., T.S.K., A.J.L. designed research, M.M.W., S.W., R.R., E.J., J.C.K., P.K., A.L., F.L., C.M., B.M., B.,M., T.B.T.A/P.T.B.H, K.L.T. performed research, M.M.W., L.B., A.J.L. analyzed data, and M.M.W., L.B., A.J.L. wrote the paper with contributions from all authors.

## SI Material

### SI Figures

**Figure S1:**
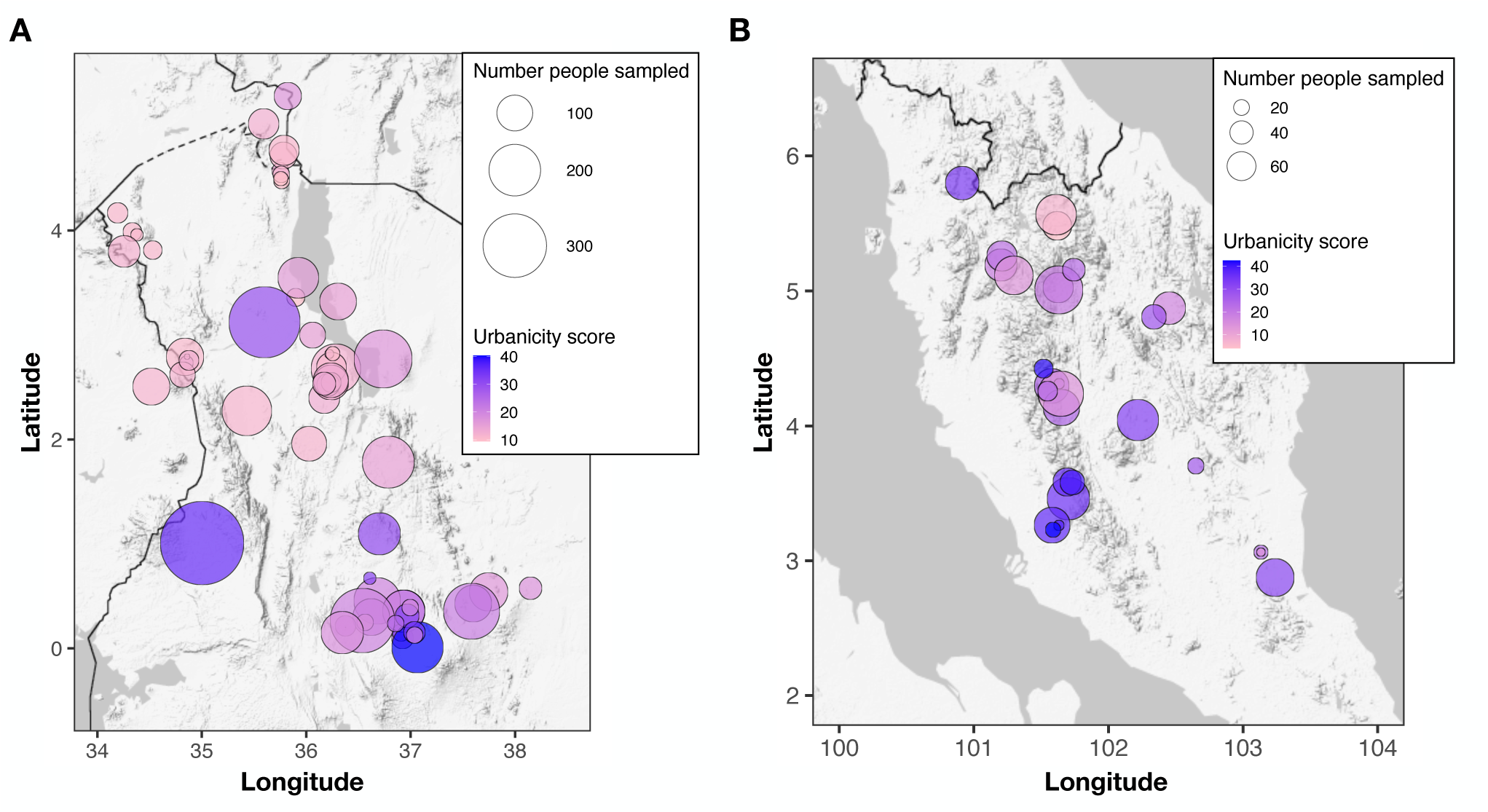
Urbanicity index score per sampling location in (A) Kenya and (B) Malaysia.

**Figure S2:**
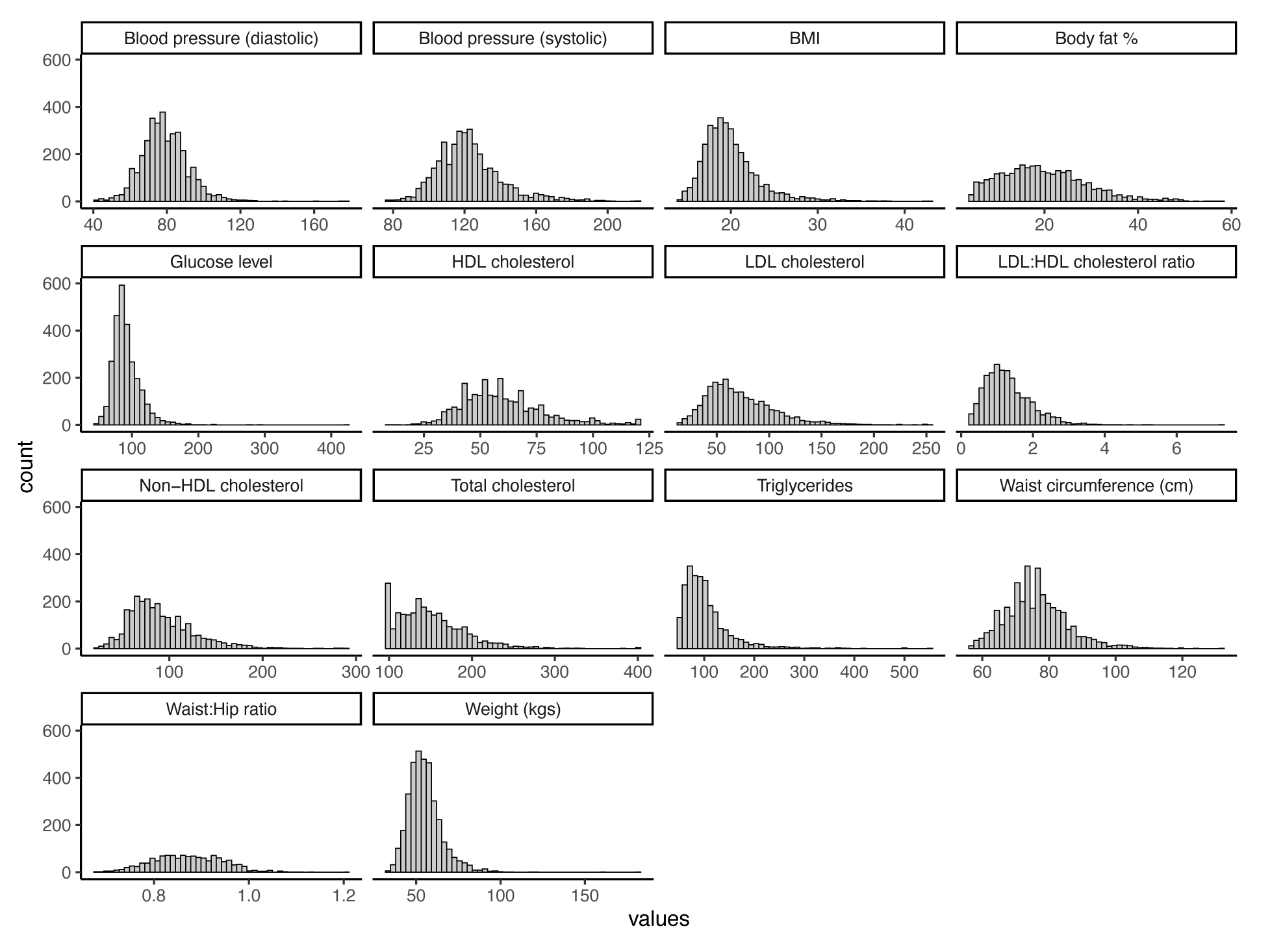
Distributions of the 14 continuous cardiometabolic biomarkers among Turkana.

**Figure S3:**
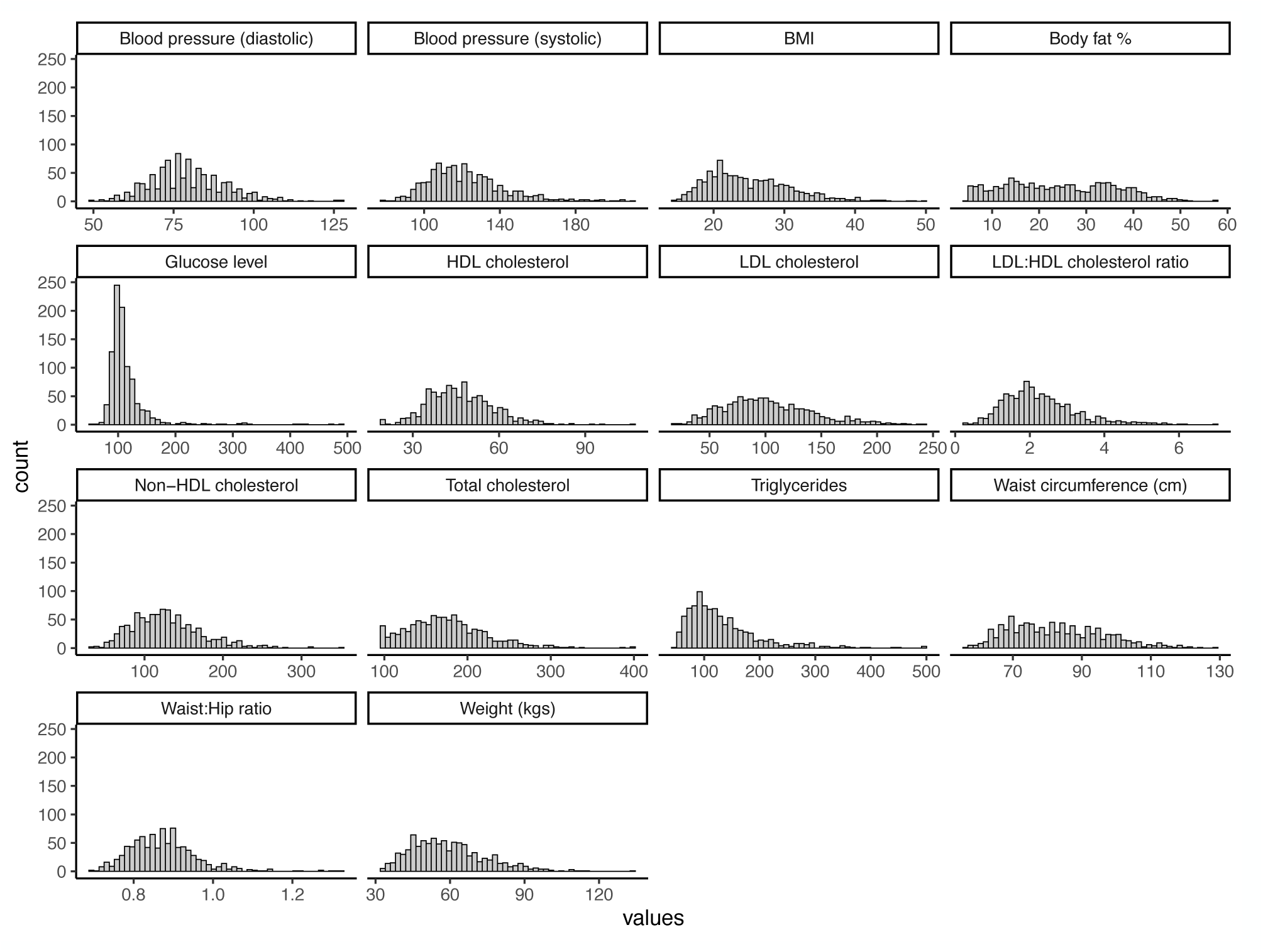
Distributions of the 14 continuous cardiometabolic biomarkers among Orang Asli.

**Figure S4:**
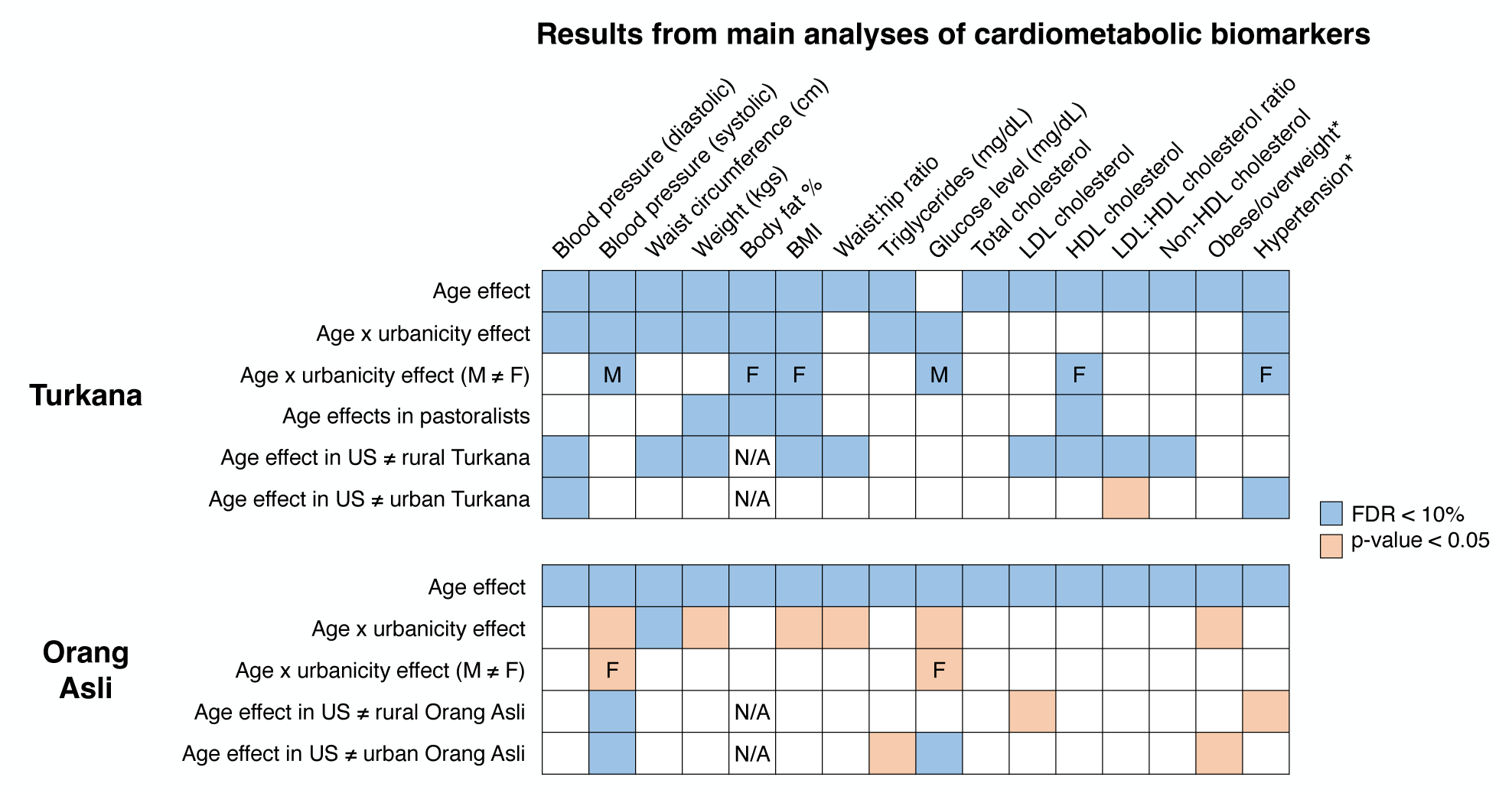
Results for each of the main analyses in Turkana and Orang Asli. Shaded boxes represent significant results at the thresholds indicated by the legend.

**Figure S5:**
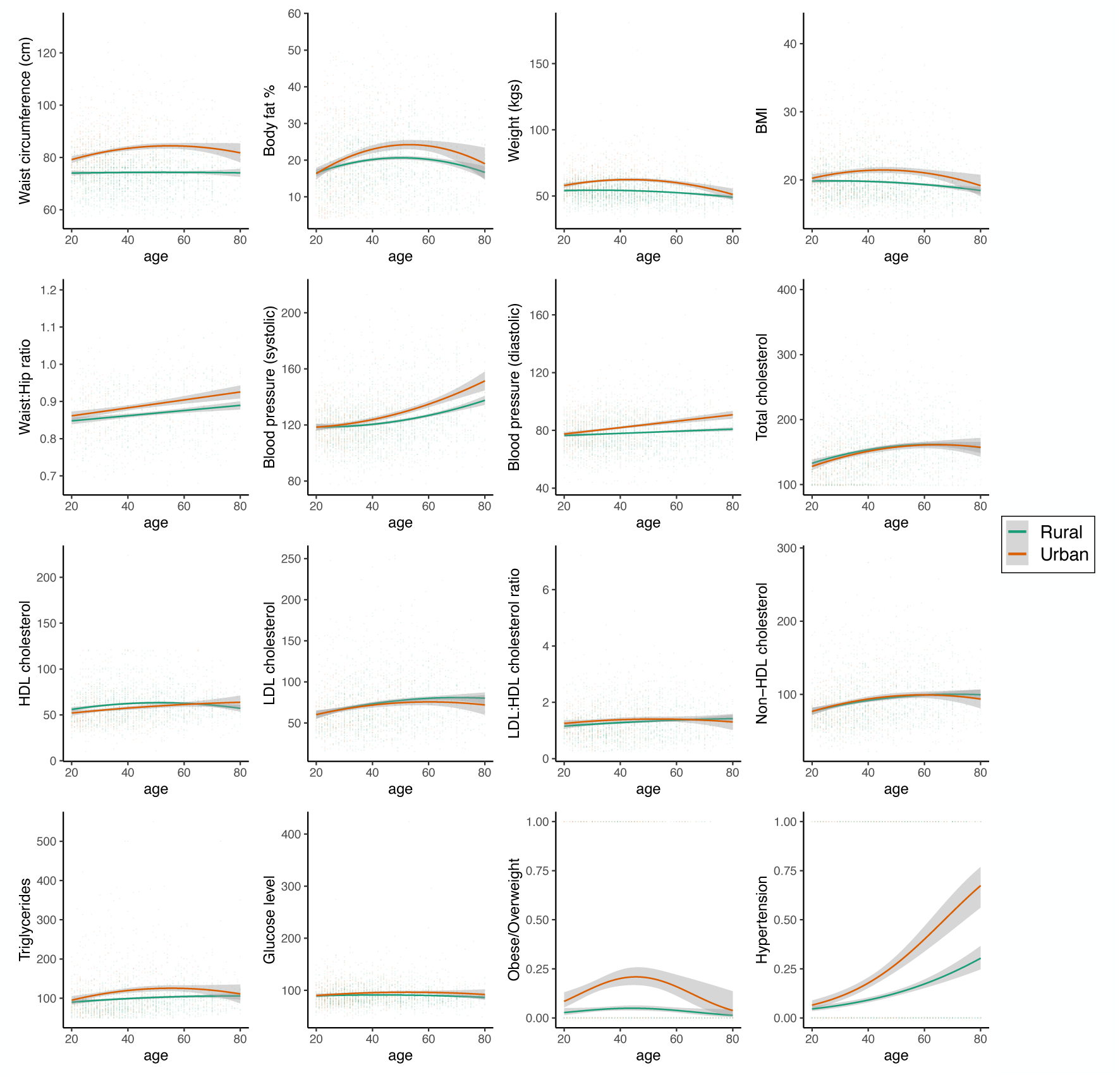
Plots of lifestyle effects on age for all continuous cardiometabolic biomarkers in Turkana. Statistical models tested urbanicity using a continuous variable; colors here are to show age trajectories for rural (green) and urban (orange) Turkana. Whether trajectory is plotted as linear or quadratic is based on our model selection tests for Turkana.

**Figure S6:**
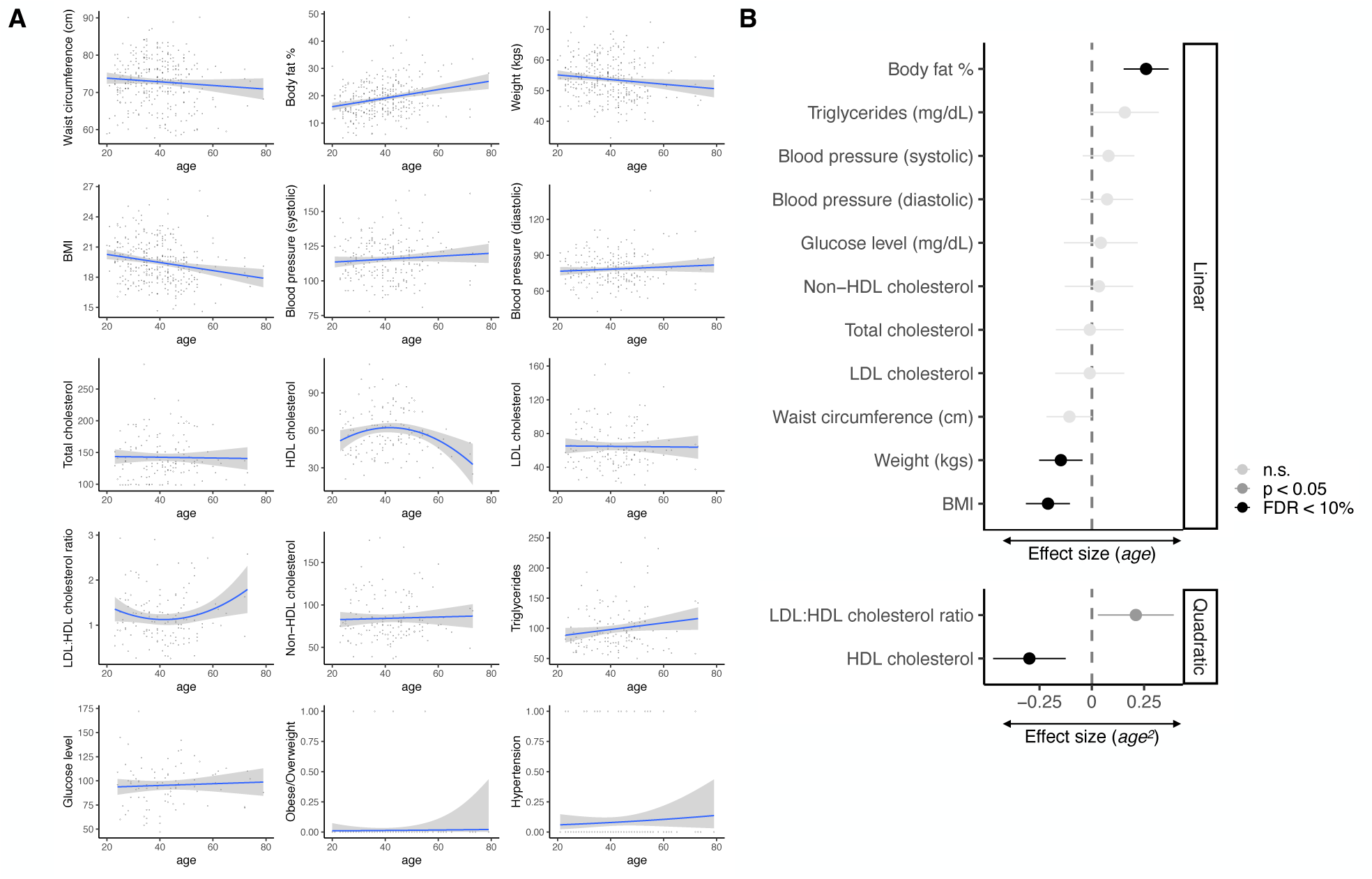
Age effects in Turkana practicing traditional, subsistence-level pastoralist lifestyles. (A) Age-associated trends for all biomarkers. (B) Standardized effect sizes of age or age^2^ (from a model selection within pastoralists).

**Figure S7:**
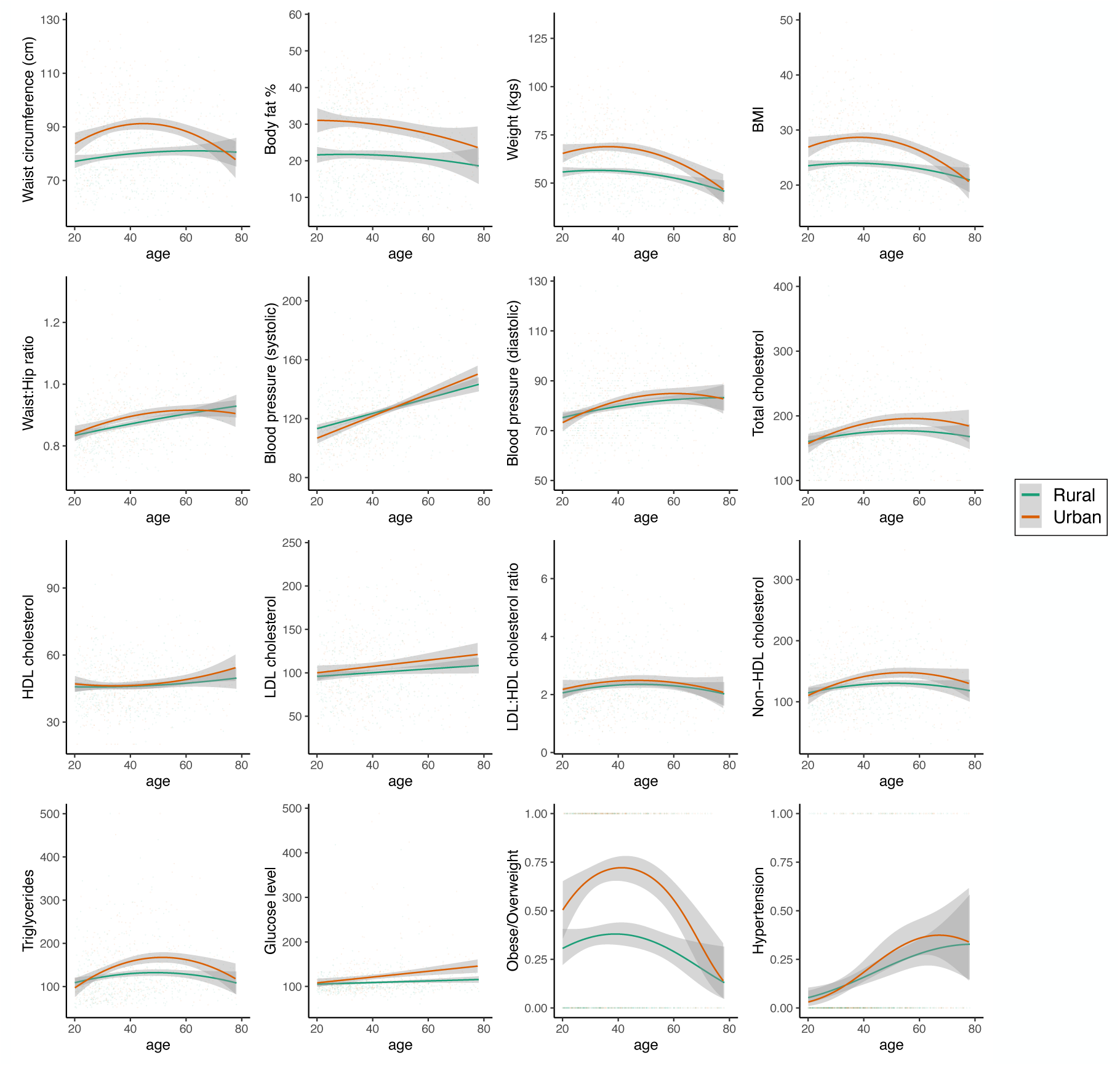
Plots of lifestyle effects on age for continuous cardiometabolic biomarkers in Orang Asli. Statistical models tested urbanicity using a continuous variable; colors here are to show age trajectories for rural (green) and urban (orange) Orang Asli. Whether trajectory is plotted as linear or quadratic is based on our model selection tests for Orang Asli.

**Figure S8:**
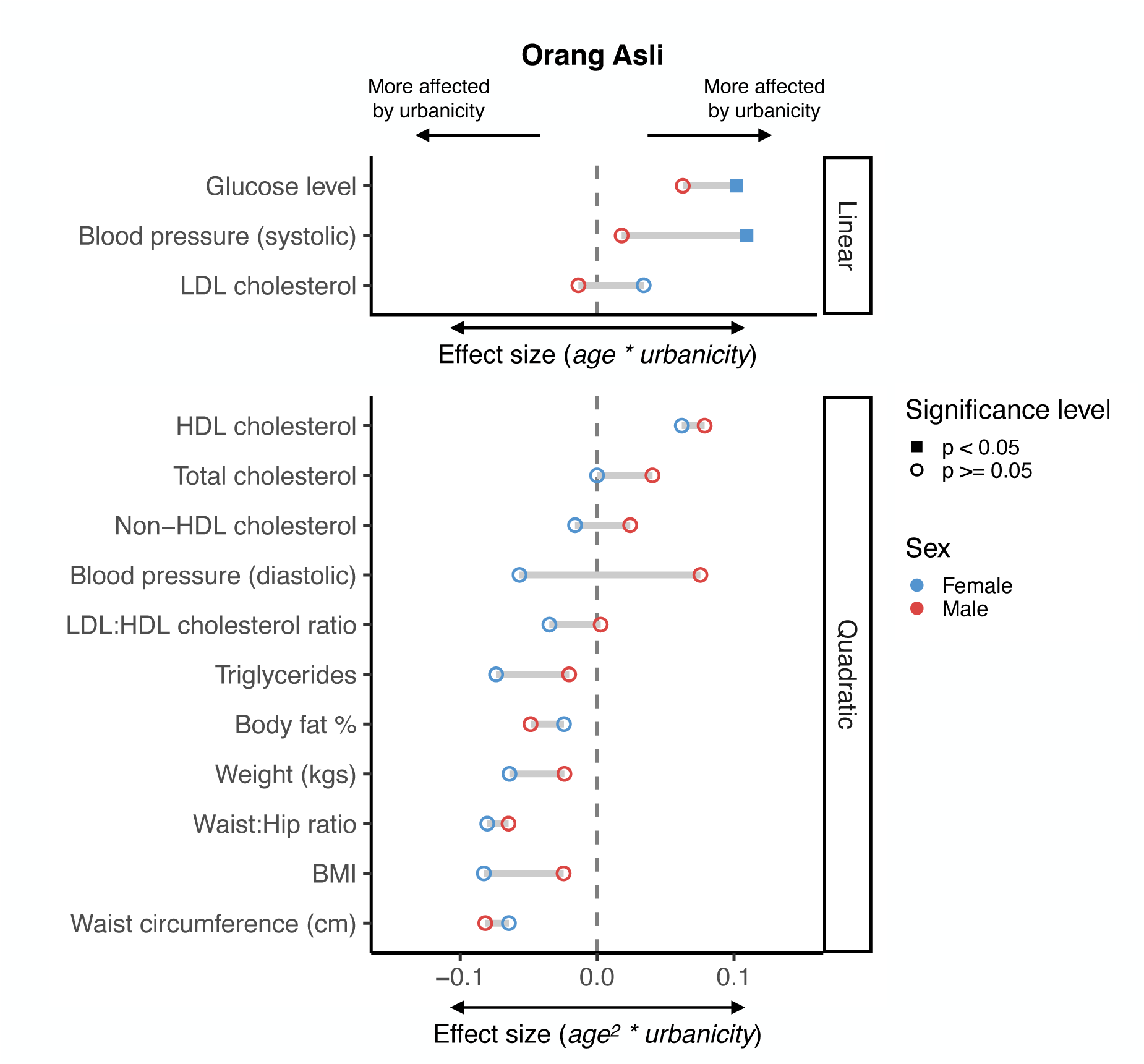
(A) Interactive age x lifestyle (or age^2^ x lifestyle) effects from models of females (blue) or males (red) in Orang Asli. Points further from the x = 0 line indicate greater lifestyle moderation of age effects. Results from categorical outcomes are not shown but are reported in Table S6.

**Figure S9:**
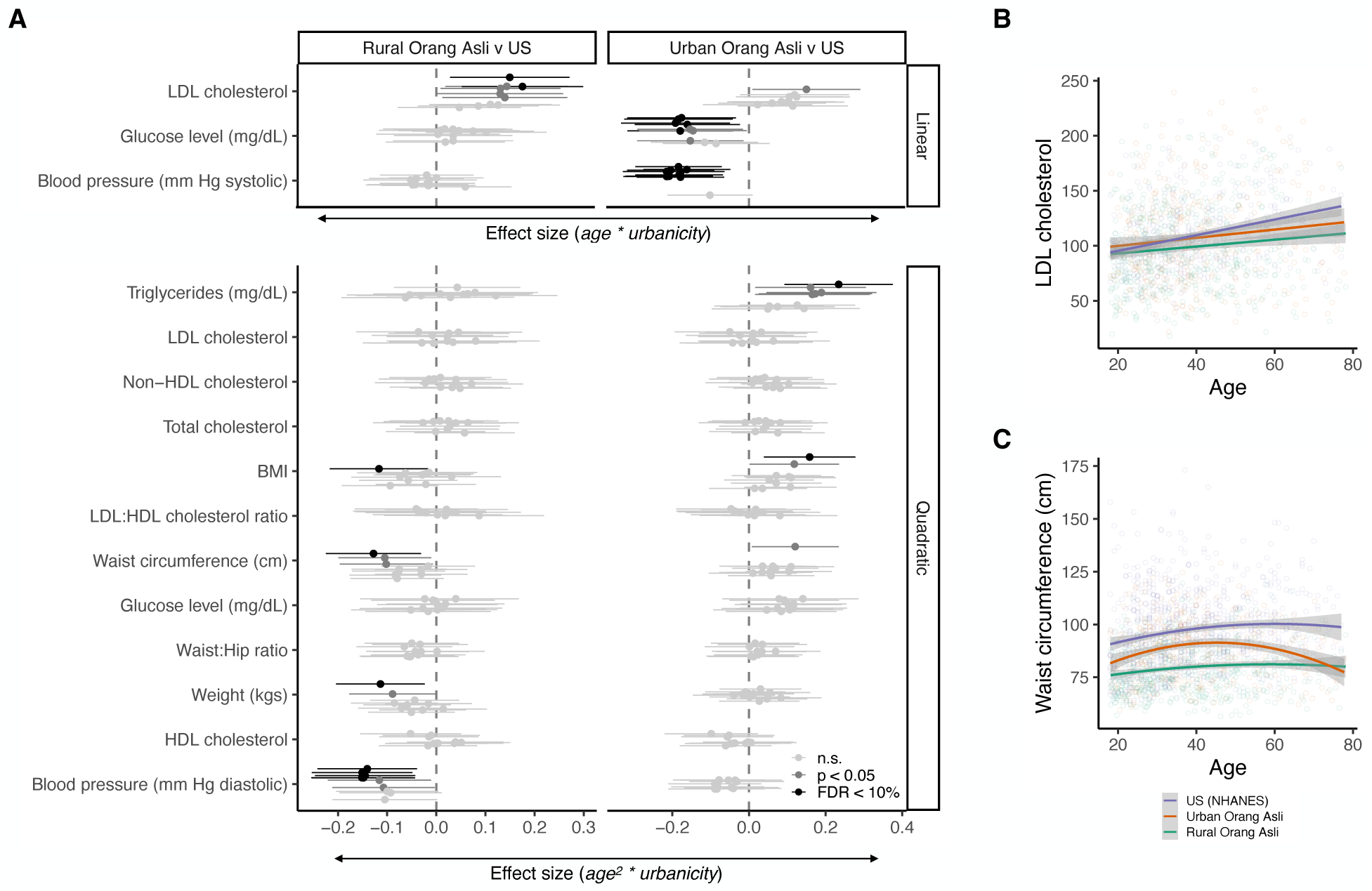
(A) Results of age x lifestyle effects for comparisons between rural Orang Asli and NHANES and urban Orang Asli and NHANES. Example plots of age-associated differences between the three lifestyle groups for biomarkers best predicted by a (B) linear and (C) quadratic age term.

**Figure S10:**
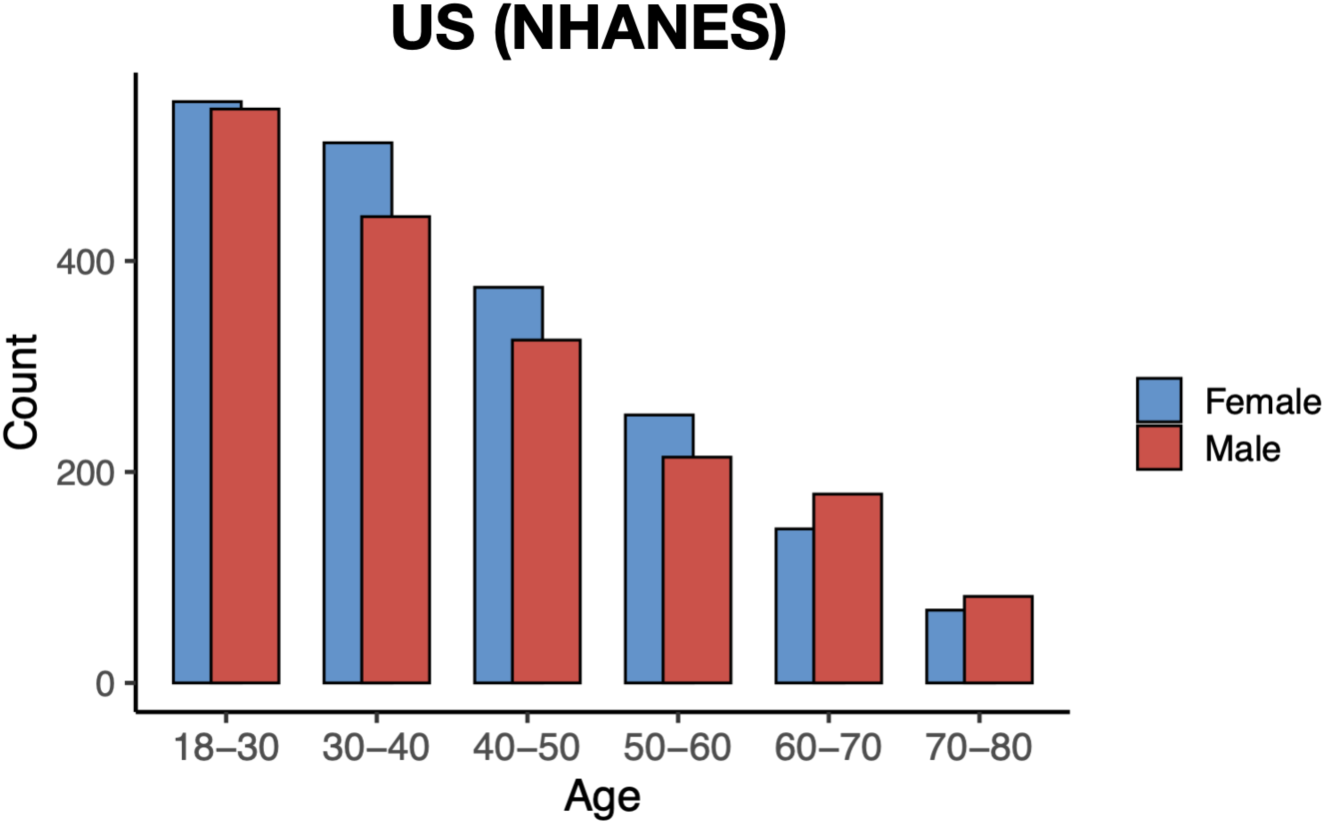
Age distribution of NHANES dataset.

**Figure S11:**
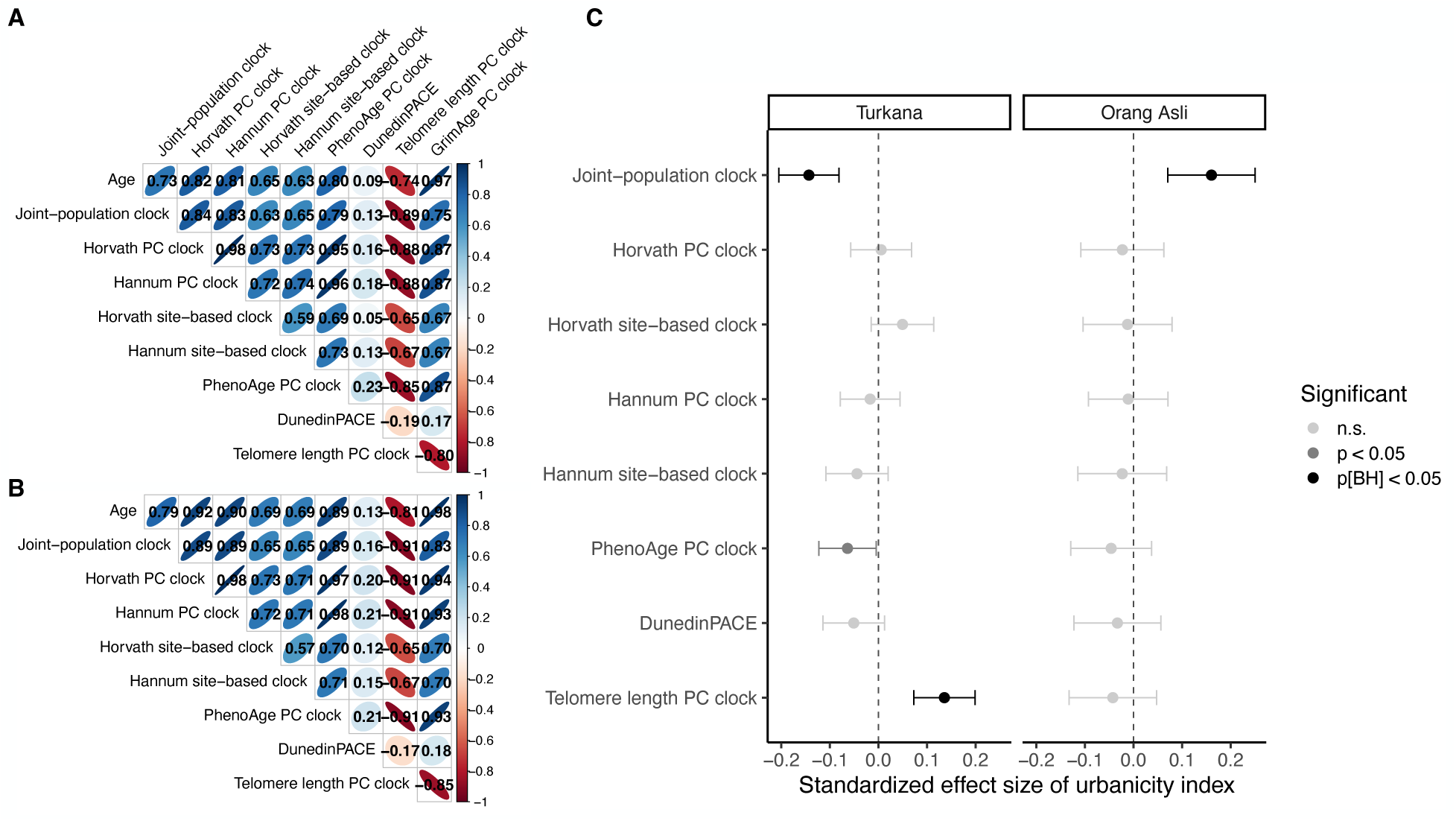
Correlogram of chronological age and biological age estimates from epigenetic clocks for Turkana. (A) and Orang Asli (B). (C) Standardized effect sizes of the urbanicity index from models predicting epigenetic age acceleration from the joint-population clock trained on DNA methylation data from Turkana and Orang Asli, the Horvath, Hannum, PhenoAge, and telomere length PC-based clocks, the Horvath and Hannum CpG site-level clocks, and DunedinPACE.

**Figure S12:**
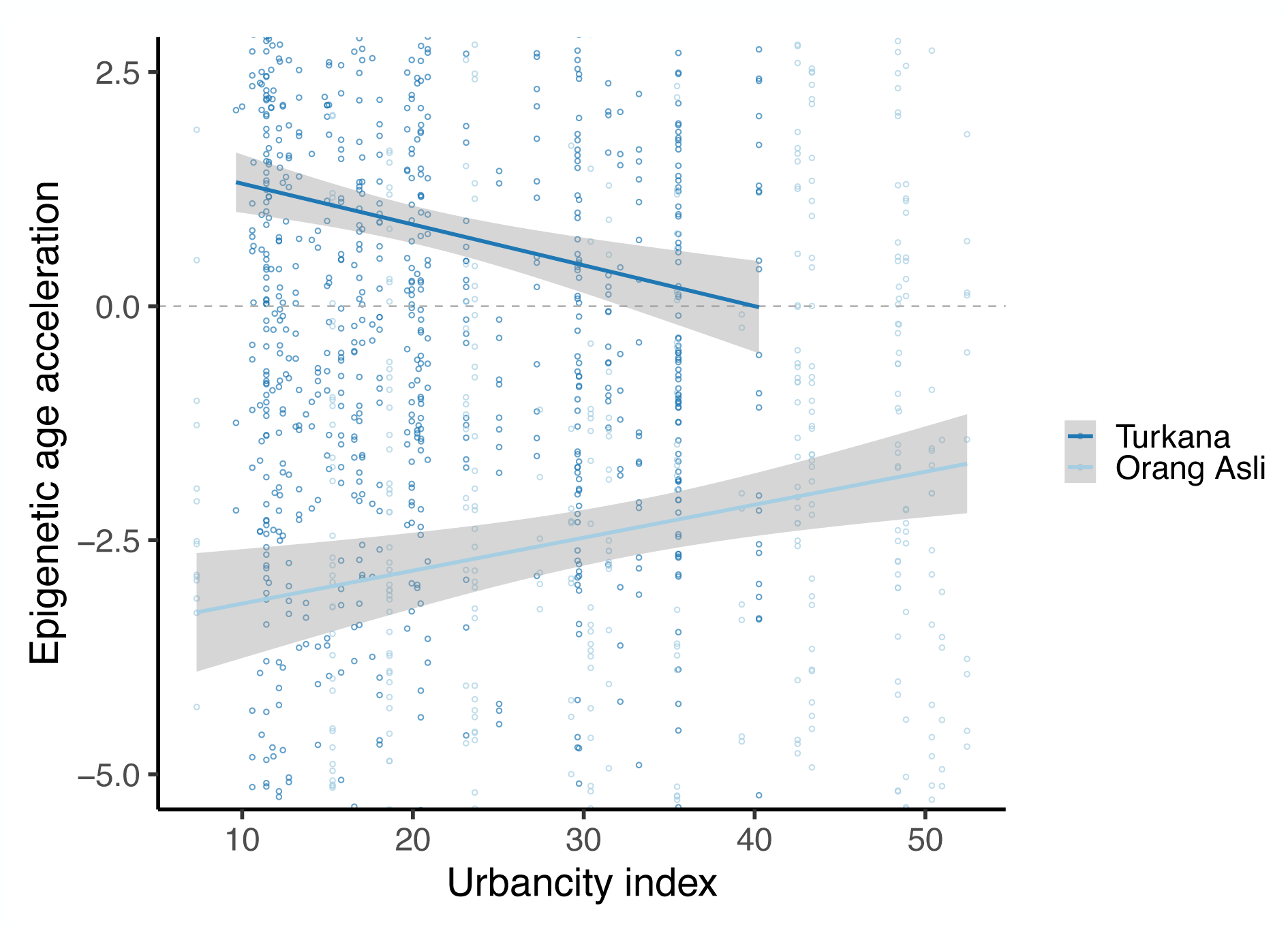
Relationship between urbanicity index and joint-population clock epigenetic age acceleration without population mean-centering.

### SI Tables

1. Demographic tables for both populations
2. Within-Turkana age, urbanicity, age x urbanicity model results
3. Within-OA age, urbanicity, age x urbanicity model results
4. Pastoralist age model results
5. Turkana sex-dependent model results
6. OA sex-dependent model results
7. OA v NHANES results (rural v NHANES, urban v NHANES)
8. Turkana vs NHANES (rural v NHANES, urban v NHANES)
9. Clock results for both groups
10. Ranges for continuous biomarker values for each dataset

## SI Results

### Sex dependent age by lifestyle effects in Orang Asli

We investigated whether age x lifestyle effects differed between males and females in Orang Asli using the same methods as for Turkana. Among Orang Asli, we were less powered to detect interactive sex-dependent effects and we did not observe any sex-dependent age x lifestyle effects that passed our stringent threshold of an FDR < 10% in one sex and p ≥ 0.05 in the other. However, we did observe some biomarkers trending towards sex-dependent age x lifestyle effects (Figure S8, Table S6).

Many prior studies in industrialized contexts have observed sex-differences in cardiometabolic traits during aging, for example, BMI (12), body fat (104, 105), and blood pressure (62, 106, 107), leading to biological hypotheses that hormones or genetic effects drive sex-dependent differences in the aging process (107–109). However, since previous work has focused on high-income, industrialized contexts, sex differences may instead reflect sex-based social roles or behaviors across these contexts. We observed few sex-based differences in how urbanicity moderated age effects and little similarity between populations, and this variability suggests that cardiometabolic aging trends may be highly dependent on local lifestyle or social features in addition to biological differences in how females and males respond to environmental stimuli.

Sex is well-established to influences social roles, labor division, and activity levels in subsistence-level societies (26, 110) and industrialized environments (111, 112), and there is increasing evidence that sex and gender roles also affect individuals’ activity levels throughout the urbanicity transition (113–115), which may interact with age. To fully disentangle these interactions between age, sex, and lifestyle, future work must leverage fine-grained activity data from females and males across a range of ages within populations undergoing lifestyle transitions. Additionally, detailed anthropological investigations are needed to illuminate how cultural expectations influence sex-dependent access to infection risk, health care, and quality of food and how these expectations change with acculturation and market-integration.

### Urban lifestyles in industrializing countries produce comparable age effects on cardiometabolic health as post-industrialized lifestyles

We wanted to understand whether post-industrialized US lifestyles exacerbated age effects to an even greater extent than we observed from our within-group analyses. We first tested this in our larger Turkana dataset, which is described in the main text. Interestingly, counter to most other biomarkers, we observed that Turkana living in urban areas had greater age-associated variance in blood pressure compared to US individuals. We also observed that Turkana living in both rural and urban areas had higher mean blood pressure than US individuals. This may be driven by the observation that individuals of East African ancestry generally have higher blood pressure than individuals of European ancestry or that blood pressure levels in recently industrializing countries in sub-Saharan Africa, Southeast Asia, and Oceania has substantially risen since the 1970s while dropping in high-income countries during this time (116). Together, our findings add to others to suggest that urbanization, and perhaps specifically lifestyle features that change relatively early in the industrialization transition, may have particularly impactful influences on blood pressure, both on baseline blood pressure levels and on how blood pressure varies with age. Combined with increasing evidence that urbanization is driving higher rates of hypertension across sub-Saharan Africa and many industrializing nations (116), mitigation strategies must be implemented quickly and efficiently to mitigate the negative effects of urbanization on blood pressure.

Using the same methods as in the Turkana dataset, we compared age x lifestyle effects between rural Orang Asli to an age and sex-matched subset of US individuals in the NHANES dataset (n = 952) for 15 biomarkers (all but body fat) and performed the same analysis comparing urban Orang Asli to US individuals. We modeled interactive age effects using the age term that best fit the data from the within-Orang Asli analyses. As in the Turkana, we performed each comparison ten times with different permutations of the NHANES data.

Comparing rural Orang Asli to the US cohort, we observed evidence for many age x lifestyle effects, though only diastolic blood pressure passed our stringent threshold for 5 of 10 permutations showing significant interactive effects at an FDR < 10% (Figure S9; Table S7). At a more liberal threshold, we observed evidence that lifestyle moderates age-related differences in weight, waist circumference, BMI, and LDL cholesterol between rural Orang Asli and the US, though many other biomarkers trended in directions we expected (Figure S9; Table S7).

Comparing urban Orang Asli to the US cohort, we observed that age-associations were steeper for urban Orang Asli for blood glucose and systolic blood pressure, and that triglycerides and BMI also trended towards being more perturbed with age in urban Orang Asli than US individuals (Figure S9; Table S7). We expect that interactive effects between rural and urban Orang Asli and US individuals are not as prevalent as those in Turkana for two primary reasons. First, the Orang Asli dataset was smaller and therefore more lowly powered to detect interactive effects. Secondly, Orang Asli, even at very low levels of urbanicity, have access to market-derived foods and also show higher average levels of adiposity, waist circumference, BMI, and blood lipids than Turkana. That urban Orang Asli exhibit similar age effects as US individuals for many biomarkers, or more exacerbated age effects for select biomarkers, suggests that the urbanicity transition that has occurred in several generations within this population is enough to recapitulate age-associated trends observed in a high-income, post-industrial country.

### Established DNA methylation clocks show few effects of urbanicity in Turkana and Orang Asli

In addition to the joint-population epigenetic clock we trained to predict age from methylation in our dataset, we investigated the effects of urbanicity on aging in both Turkana and Orang Asli from 10 previously validated epigenetic clocks. These included first-generation clocks trained on chronological age, and second– and third-generation clocks trained on pace of aging or molecular or phenotypic aging (clocks detailed in Methods). In Turkana, epigenetic age acceleration was negatively associated with higher urbanicity for the joint-population clock, as reported in the Results, and positively associated with the PC telomere length clock (β = 0.13, p_BH_ = 2.06 x 10^−4^) (Figures 5B, S11, Table S9). Within Orang Asli, epigenetic age acceleration was not significantly associated with any other clock besides the joint-population clock, as reported in the Results (Figures 5B, S11; Table S9).

